# Pesticides and lifestyle factors are associated with disease severity of Parkinson’s disease: a longitudinal study

**DOI:** 10.1101/2024.09.06.24313168

**Authors:** Theresa Lüth, Amke Caliebe, Carolin Gabbert, Sebastian Sendel, Björn-Hergen Laabs, Inke R. König, Christine Klein, Joanne Trinh

## Abstract

**Objective:** To longitudinally analyze the impact of the environment and lifestyle on PD motor sign severity in LRRK2 p. Gly2019Ser-related PD (*LRRK2*-PD) and idiopathic PD (iPD).

**Background:** There is increasing evidence that the environment impacts disease severity. Recent studies have shown that pesticide exposure is associated with a faster disease progression. However, the relationship between smoking, caffeine, and disease severity has not yet been investigated longitudinally.

**Methods:** In this longitudinal study, we included patients with iPD from the PPMI Online (*N*=2815) and Fox Insight (*N*=2319) cohorts, as well as patients with *LRRK2*-PD (*N*=81) from Fox Insight. Motor signs were assessed with the MDS-UPDRS Part II, and patients were assessed multiple times, followed up to 35 months or 60 months in the PPMI Online or Fox Insight cohort, respectively. The motor sign severity over time was analyzed by applying a linear mixed effects model. The outcome was the cumulative score of the MDS-UPDRS Part II questionnaire. Subsequently, we investigated the association between environmental exposure, lifestyle factors and motor signs. Pesticide exposure in a work setting, smoking, coffee, black tea, green tea, and caffeinated soda consumption were assessed using the validated PD-RFQ-U questionnaires. The mixed effects model included the environmental and lifestyle factors as binary (yes/no) variables.

**Results:** When comparing *LRRK2*-related PD and iPD, motor signs were less severe in patients with *LRRK2-*PD compared to iPD (β=-0.23, *p*=0.005). In *LRRK2*-PD, black tea consumption was associated with less severe motor signs (β=-0.51, *p*=0.028). In patients with iPD, we observed that pesticide exposure was associated with more severe motor signs over time in PPMI-Online (β=0.23, *p*=3.56×10^-9^). Smoking was associated with a higher motor signs score in PPMI-Online (β=0.13, *p*=0.001). Lastly, caffeinated soda was associated with more severe motor signs in patients with iPD from PPMI-Online (β=0.15, *p*=3.84×10^-8^) and Fox Insight (β=0.09, *p*=0.031).

**Conclusions:** Our results provide further evidence of the importance of environment and lifestyle in PD, even after the disease onset. We suggest that pesticide exposure and lifestyle factors may affect disease severity in patients with *LRRK2*-PD and iPD; still, further validation is necessary.

## 1. Introduction

PD is a progressive neurodegenerative disorder that currently affects close to 12 million patients worldwide (Collaborators, 2024; Dorsey et al., 2018). From 1990 to 2021, the age-standardized number of individuals with PD increased by 60.7%; at the same time, the number of Alzheimer’s disease patients increased only by 3.2% (Collaborators, 2024). As the number of cases has risen constantly over the last decade, PD will be a large burden to society, especially since there is no cure available and only medication to manage the signs and symptoms of the disease. Among the ∼15% of PD patients where the disorder is explained by a single pathogenic variant (i.e., monogenic PD) or strong risk factor (Cook et al., 2024; Westenberger et al., 2024), the LRRK2 G2019S variant is the most frequent cause of autosomal-dominant PD (*LRRK2*-PD). Recently, a 3.5-year longitudinal study assessed disease severity in patients with *LRRK2*-PD in the 23andMe dataset, and it has been suggested that patients present predominantly with PD motor feature impairment and have less severe non-motor features compared to patients with idiopathic PD (iPD) (Kmiecik et al., 2024).

Besides genetics, the environment and lifestyle are of great importance in PD. Pesticide exposure is the strongest environmental risk factor for PD (Dorsey and Bloem, 2024; Noyce et al., 2012) and is associated with an earlier age at onset (AAO) (Gamache et al., 2019; Ratner et al., 2014). Additionally, there has been recent evidence that pesticides impact PD after disease onset, as a longitudinal study showed that exposure is associated with faster disease progression (Brown et al., 2024; Li et al., 2023). Smoking and coffee are the most crucial lifestyle factors associated with reduced PD risk (Noyce et al., 2012) and are analogously associated with later AAO, as demonstrated by many studies (Gabbert et al., 2022; Gigante et al., 2018; Gigante et al., 2017; Luth et al., 2020; Noyce et al., 2012; Wijeyekoon et al., 2017). With regard to monogenic *LRRK2*-PD, smoking, coffee and black tea consumption are associated with a later AAO as well (Luth et al., 2020; Yahalom et al., 2020). Interestingly, caffeinated soda, on the other hand, was found to be associated with earlier AAO in *LRRK2*-PD (Luth et al., 2020).

Thus far, the relationship between smoking, caffeine, and disease severity has not been investigated longitudinally. In contrast to the proposed protective effect of smoking on PD susceptibility and onset, a cross-sectional assessment suggested an association between smoking and more severe motor and non-motor features in PD (Gabbert et al., 2023). This observation highlights that different mechanisms might mediate the impact of environment and lifestyle before and after disease onset. Thus, a more thorough and longitudinal analysis is required to elucidate environmental and lifestyle factors’ association with PD severity over time. Results from longitudinal cohort studies provide a higher evidence level when compared to case-control studies, which suffer from recall bias, retrospective study design, enhanced confounding and lack of differentiation between pre- or post-disease exposures. In addition, distinguishing between *LRRK2*-PD and iPD will allude to a better understanding of the natural history of the different PD subtypes and possibly also the specific treatment responses.

Large available PD cohorts, including multiple patient assessments over the years, are an essential resource for investigating disease severity over time. Fox Insight is one of the largest cohorts of such kind, and participants are followed up for five years (Gottesman et al., 2024), as well as Parkinson’s Progression Markers Initiative Online Study (PPMI Online), in which patients are followed up for three years (Giakas et al., 2022; Marek et al., 2018).

Therefore, we investigated motor signs longitudinally in patients with *LRRK2*-PD from Fox Insight and patients with iPD from PPMI Online and Fox Insight. Herein, we focused on the association of environmental and lifestyle factors (i.e., pesticide exposure, smoking and caffeine consumption) and motor sign severity over time.

## 2. Methods

### 2.1 Longitudinal Cohorts and Participants Demographics

The online cohorts Fox Insight and PPMI Online are focused on PD research, and both provide a platform for patients to participate. The Fox Insight data consists of up to five years of routine longitudinal assessments (health and medical questionnaires) and additional one-time questionnaires about, e.g., environmental exposure and lifestyle habits assessed at enrollment (Gottesman et al., 2024). The PPMI Online study is also an online cohort, recruiting participants with and without PD. It is linked to the PPMI study and includes longitudinal assessments of motor and non-motor features of PD and environmental/lifestyle data assessed at different study time points (Giakas et al., 2022; Marek et al., 2018).

The participants were filtered for AAO >25 years, disease duration <50 years and at least three longitudinal assessments of motor signs. This data harmonization step was included to ensure an unbiased approach without extreme outliers for a fair outcome of the statistical analysis and to obtain an approximately equal number of patients per assessment. We included *N*=2815 patients with iPD from the PPMI Online cohort and *N*=2319 from Fox Insight. In addition, we included *N*=81 patients with *LRRK2*-PD from Fox Insight (**Figure 1**). In this study, we focused on PD motor features as those were assessed with the same questionnaire in both cohorts, and it has been reported previously that *LRRK2*-PD patients are predominantly affected by motor impairments (Kmiecik et al., 2024).

**Figure 1.**
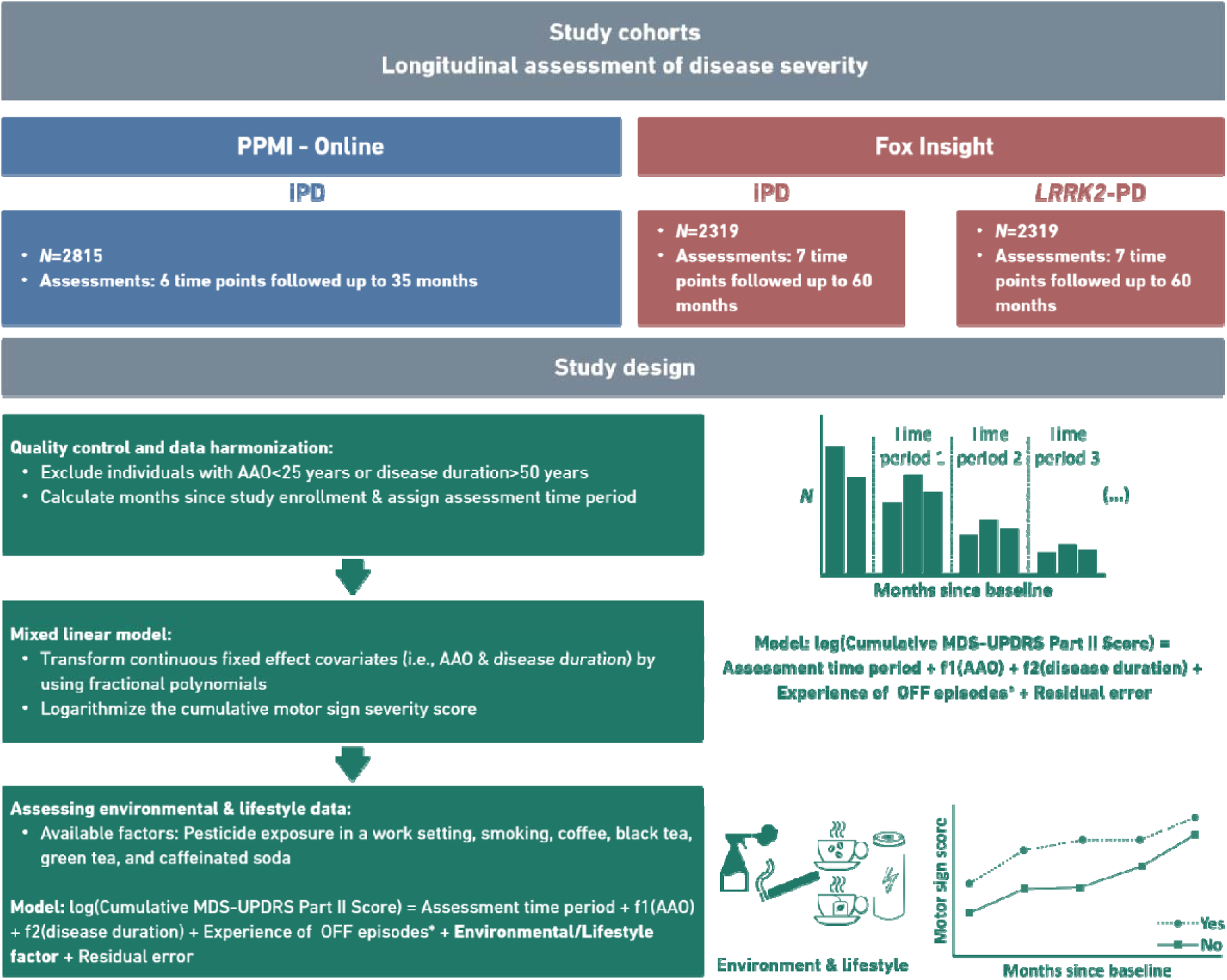
Study overview. The diagram illustrates the included longitudinal cohorts and the study design (data quality control, construction of the statistical model and overlaying of environmental/lifestyle data). *N*=number of individuals, iPD=Patients with idiopathic Parkinson’s disease, and *LRRK2*-PD=Patients with PD who carry the LRRK2 p.Gly2019Ser variant, f1/f2=polynomial transformation age at onset or disease duration. *If applicable

In patients with iPD, there was a similar AAO (PPMI Online: mean AAO (±*SD*)=63.55 years (±8.93), Fox Insight: mean AAO (±*SD*)=61.57 years (±8.83)) and disease duration (PPMI Online: mean disease duration (±*SD*)=4.18 years (±4.08), Fox Insight: mean disease duration (±*SD*)=4.38 years (±4.17)) at baseline in both cohorts. However, patients with *LRRK2*-PD had a slightly younger AAO (mean AAO (±*SD*)= 60.06 years (±9.02)) and longer disease duration (mean disease duration (±*SD*)=5.52 years (±4.53)) (**Table 1**). The severity of motor signs was assessed using the MDS-UPDRS Part II score. The individual scores for PD motor features included in the questionnaire were added to obtain the cumulative MDS-UPDRS Part II score.

**Table 1.** Demographic overview of the longitudinal PD cohorts.

|  | PPMI - Online | Fox Insight |  |
| --- | --- | --- | --- |
|  | iPD | iPD | *LRRK2*-PD |
| ***N*** | 2815 | 2319 | 81 |
| **Number of men (%)** | 1690 (60.0%) | 1247 (53.8%) | 42 (51.9%) |
| **Mean AAE ($\pm SD$)** | 67.73 years ($\pm 8.29$) | 65.95 years ($\pm 8.12$) | 65.08 years ($\pm 8.05$) |
| **Mean AAO ($\pm SD$)** | 63.55 years ($\pm 8.93$) | 61.57 years ($\pm 8.83$) | 60.06 years ($\pm 9.02$) |
| **Mean disease duration at baseline ($\pm SD$)** | 4.18 years ($\pm 4.08$) | 4.38 years ($\pm 4.17$) | 5.52 years ($\pm 4.53$) |
| **Mean study enrollment time ($\pm SD$)** | 28.32 months ($\pm 1.40$) | 52.54 months ($\pm 2.52$) | 52.49 months ($\pm 2.48$) |
| **Mean motor signs progression ($\pm SD$)** | 2.06 ($\pm 4.81$) | 3.23 ($\pm 6.14$) | 2.93 ($\pm 5.38$) |

*N*=Number of individuals, AAE=Age at examination, AAO=Age at onset, iPD=Patients with idiopathic Parkinson's disease, *LRRK2*-PD=Patients with PD that carry the *LRRK2* p.Gly2019Ser variant, Motor signs progression= (Cumulative MDS-UPDRS Part II score at the last assessment time period) - (Cumulative MDS-UPDRS Part II score at baseline)

Participants of the PPMI Online cohort were followed up for up to 35 months (mean study enrollment time (±*SD*): 28.32 months (±1.40)) and participants of the Fox Insight cohort for up to 60 months (iPD: mean study enrollment time (±*SD*)=52.54 months (±2.52), *LRRK2*-PD: mean study enrollment time (±*SD*)=52.49 months (±2.48)). By calculating the months since the baseline (at 0 months) of the individual assessments, we assigned them to one of either six or seven different assessment time periods in the PPMI-Online or Fox Insight data sets, respectively. The 35-month enrollment study time of the PPMI-Online cohort was divided into six to nine-month-long time periods, depending on the number of observations per time period (**Supplementary Figure 1A**). The 60-month enrollment study time of the Fox Insight cohort was divided into 10-month-long time periods (**Supplementary Figure 1B and 1C**). In the rare case that a patient was assessed two times per assigned time period, we included only the first assessment in our analysis.

### 2.2 Environmental and Lifestyle Factors

The PD Risk Factor Questionnaire (PD RFQ-U) was used to assess environmental and lifestyle information in the PPMI Online and Fox Insight cohort. The National Institute of Environmental Health Sciences constructed and validated the questionnaire for interview and self-reporting settings (https://www.commondataelements.ninds.nih.gov/report-viewer/23723/Risk%20Factor%20Questionnaire%20(RFQ-U)%20-%20Pesticides%20(Work)). In the PD RFQ-U, pesticide exposure in a work setting is defined as being ever exposed to pesticides in your job; smoking is defined as smoking more than 100 cigarettes in your lifetime and caffeine consumption is defined as drinking coffee, black tea, green tea or caffeinated soda at least once per week for more than six months.

### 2.3 Linear Mixed Effects Model

To evaluate the longitudinal data statistically, we applied linear mixed effects models. The model’s outcome was the log-transformed cumulative MDS-UPDRS Part II score (range 0-52) to achieve a more appropriate normal distribution of the outcome variable (**Supplementary Figure 2**). We included time (six or seven time periods, categorical variable), disease duration (in years), AAO, (in years) and experience of OFF episodes (binary yes/no variable) as fixed effects and the patient ID as the random effect in the model. The continuous fixed effect covariates (i.e., AAO & disease duration) were transformed using fractional polynomials to achieve a more linear correlation with the outcome variable (**Supplementary Figure 3**). The information about the experience of OFF episodes was only available for the Fox Insight cohort (**Figure 1**). In Fox Insight, we also applied the mixed linear effect model to the complete data set, including patients with iPD together with *LRRK2*-PD patients into one model and the *LRRK2*-PD/iPD status as a binary variable as an additional fixed effect.

Linear mixed effects models were also applied to explore the association between environmental exposures, lifestyle factors and disease severity over time. The individual environmental and lifestyle factors were included as fixed effects in the form of binary variables (yes/no).

The analyses were performed in R (v.4.3.1) (R Core Team, 2023) with the *lme4* (v.1.1-34)and *lmerTest* (v.3.1-3) packages (Bates et al., 2015; Kuznetsova et al., 2017). The continuous fixed-effect covariates were transformed with a multiple fractional polynomials (*mfp*) function from the *mfp* (v. 1.5.4) package (Ambler, 2023). The analysis of the association between pesticide exposure and motor sign severity was interpreted for significance, based on the presence of the ‘a priori’ hypothesis on the association between pesticide and PD severity. The association with pesticides was tested for significance in iPD patients from PPMI-Online, Fox Insight and *LRRK2*-PD patients from Fox Insight. Thus, the significance level was adjusted for performing three tests to α = 0.05/3 = 0.017. All other analyses in this study were exploratory and *p*-values were not corrected for multiple testing.

## 3. Results

### 3.1 Disease Severity Over Time in iPD and LRRK2-PD

We included patients with iPD from the PPMI Online cohort. The mean enrollment time was 28.32 months, and the mean increase in motor features, as assessed with the MDS-UPDRS Part II, was 2.06. During the six assessment time periods, a gradual increase in motor sign severity was observed (**Figure 2**). The gradual rise of motor sign severity was also illustrated with the linear mixed effects model (**Table 2**), as there was an increasing effect size for each subsequent time period, demonstrating the longitudinal increase in disease severity (**Table 2**).

**Figure 2.**
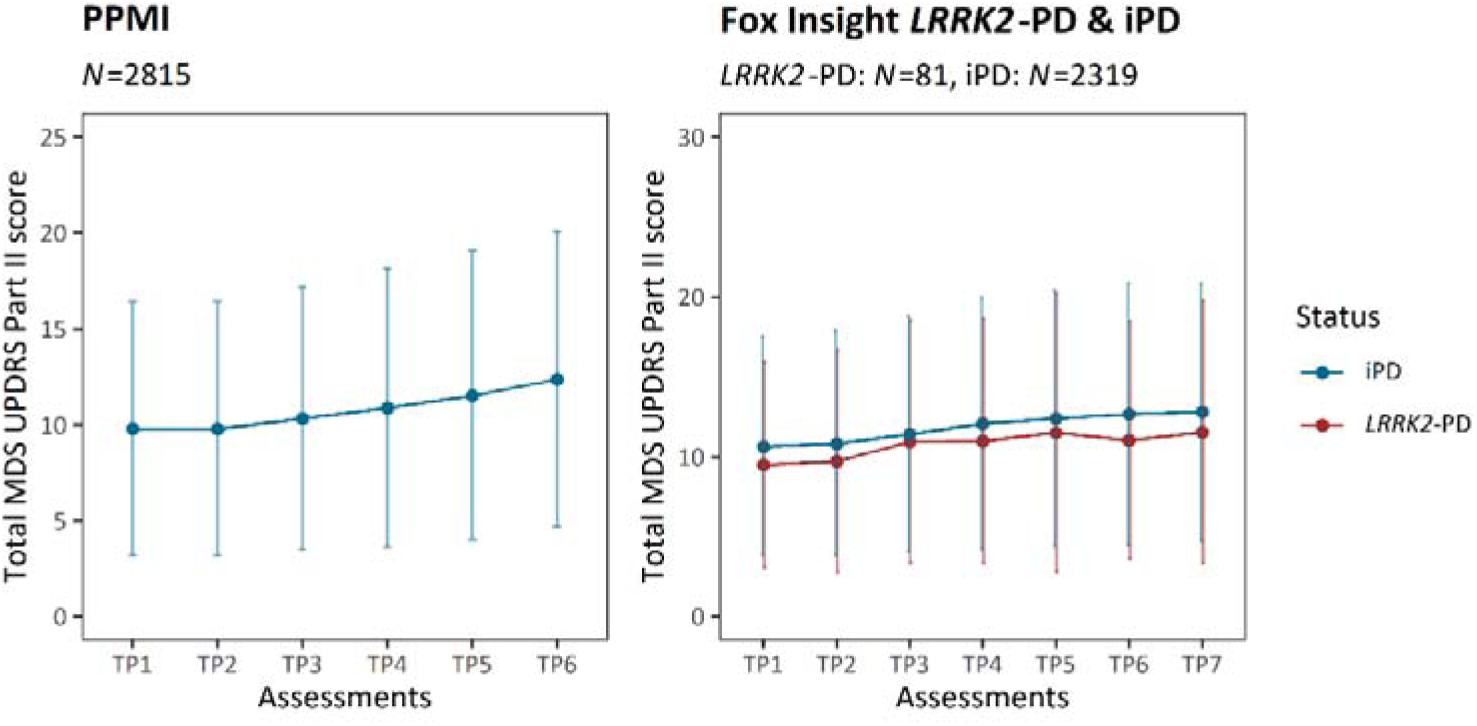
Motor sign severity over time. The plots show the progression of PD motor features along the longitudinal assessments. At each time period, the mean cumulative MDS-UPDRS Part II score is indicated, and the error bars show the corresponding standard deviation. Patients with iPD are shown in blue (PPMI and Fox Insight) and patients with *LRRK2*-PD are shown in red (Fox Insight). iPD=Patients with idiopathic Parkinson’s disease, *LRRK2*-PD=Patients with PD that carry the LRRK2 p.Gly2019Ser variant, TP=Time period, *N*=number of individuals.

**Table 2.** The motor sign severity was evaluated longitudinally over time and assessed with a linear mixed model in the PPMI and Fox Insight cohort.

|  | Estimate | SE | p-value |
| --- | --- | --- | --- |
| **PPMI – Online (iPD: N=2815)** |  |  |  |
| Time period 2 | 0.0001 | 0.01 | 0.993 |
| Time period 3 | 0.07 | 0.01 | $9.78 \times 10^{-13}$ |
| Time period 4 | 0.12 | 0.01 | $<2.00 \times 10^{-16}$ |
| Time period 5 | 0.19 | 0.01 | $<2.00 \times 10^{-16}$ |
| Time period 6 | 0.26 | 0.01 | $<2.00 \times 10^{-16}$ |
| AAO | -0.05 | 0.01 | $1.57 \times 10^{-04}$ |
| Disease duration | 0.64 | 0.03 | $<2.00 \times 10^{-16}$ |
| **Fox Insight (iPD: N=2319)** |  |  |  |
| Time period 2 | 0.01 | 0.01 | 0.197 |
| Time period 3 | 0.08 | 0.01 | $1.39 \times 10^{-11}$ |
| Time period 4 | 0.15 | 0.01 | $<2.00 \times 10^{-16}$ |
| Time period 5 | 0.21 | 0.01 | $<2.00 \times 10^{-16}$ |
| Time period 6 | 0.27 | 0.01 | $<2.00 \times 10^{-16}$ |
| Time period 7 | 0.31 | 0.01 | $<2.00 \times 10^{-16}$ |
| AAO | -0.03 | 0.01 | 0.019 |
| Disease duration | 0.71 | 0.05 | $<2.00 \times 10^{-16}$ |
| OFF-episodes | 0.25 | 0.03 | $<2.00 \times 10^{-16}$ |
| **Fox Insight (LRRK2-PD: N=81)** |  |  |  |
| Time period 2 | -0.04 | 0.06 | 0.527 |
| Time period 3 | 0.05 | 0.06 | 0.410 |
| Time period 4 | 0.17 | 0.06 | 0.007 |
| Time period 5 | 0.19 | 0.06 | 0.003 |
| Time period 6 | 0.29 | 0.07 | $4.98 \times 10^{-5}$ |
| Time period 7 | 0.36 | 0.07 | $1.38 \times 10^{-7}$ |
| AAO | 0.14 | 0.92 | 0.880 |
| Disease duration | 0.39 | 0.18 | 0.034 |
| OFF-episodes | 0.75 | 0.16 | $6.13 \times 10^{-6}$ |
| **Fox Insight (LRRK2-PD & iPD)** |  |  |  |
| Time period 2 | 0.02 | 0.01 | 0.093 |
| Time period 3 | 0.08 | 0.01 | $5.63 \times 10^{-14}$ |
| Time period 4 | 0.15 | 0.01 | $<2.00 \times 10^{-16}$ |
| Time period 5 | 0.22 | 0.01 | $<2.00 \times 10^{-16}$ |
| Time period 6 | 0.27 | 0.01 | $<2.00 \times 10^{-16}$ |
| Time period 7 | 0.32 | 0.01 | $<2.00 \times 10^{-16}$ |
| LRRK2-PD | -0.23 | 0.08 | 0.005 |
| AAO | -0.04 | 0.01 | 0.003 |
| Disease duration | 0.73 | 0.04 | $<2.00 \times 10^{-16}$ |
| OFF-episodes | 0.27 | 0.03 | $<2.00 \times 10^{-16}$ |

N=Number of individuals, iPD=Patients idiopathic Parkinson's disease, LRRK2-PD= Patients with PD that carry the LRRK2 p.Gly2019Ser variant, Formula in R (package lme4): lmer(log(Cumulative MDS-UPDRS Part II Score) ~ Assessment time periods+ mfp transformed AAO + mfp transformed disease duration + LRRK2-PD/iPD\* + Experience of OFF episodes\* + (1|Patient ID)), \*If applicable
Tested for significance at $\alpha = 0.017$
Baseline categories: Time period=Time period 1 (i.e., assessment at enrollment)

On the other hand, patients with iPD and *LRRK2*-PD were included from the Fox Insight cohort, where the patients had a mean enrollment time of 52.54 months and 52.49 months, respectively (**Table 1**). We observed more severe motor signs over all seven assessments among patients with iPD (mean motor features progression=3.23) compared to patients with *LRRK2*-PD (mean motor features progression=2.93) (**Table 1**, **Figure 2**). The less severe motor signs of patients with *LRRK2*-PD were confirmed with a linear mixed effects model (β=-0.23, *SE*=0.08, *p*=0.005) (**Table 2**).

### 3.2 Pesticide Exposure in a Work Setting

Within the PPMI Online cohort, approximately 15% (*N*=2467) of the patients with iPD were exposed to pesticides in a work setting. After the 35-month longitudinal assessment of motor signs, iPD patients who were exposed to pesticides had more severe motor signs (mean cumulative MDS-UPDRS Part II (±*SD*)=14.15 (±8.24)) compared to those who were not exposed (mean cumulative MDS-UPDRS Part II (±*SD*)=11.99 (±7.49)) (**Figure 3**). The linear mixed effects model confirmed the significant association between pesticide exposure and disease severity, including six assessment time periods throughout the study (β=0.23, *SE*=0.04, *p*=3.56×10^-9^, adjusted α = 0.017) (**Table 3**).

**Figure 3.**
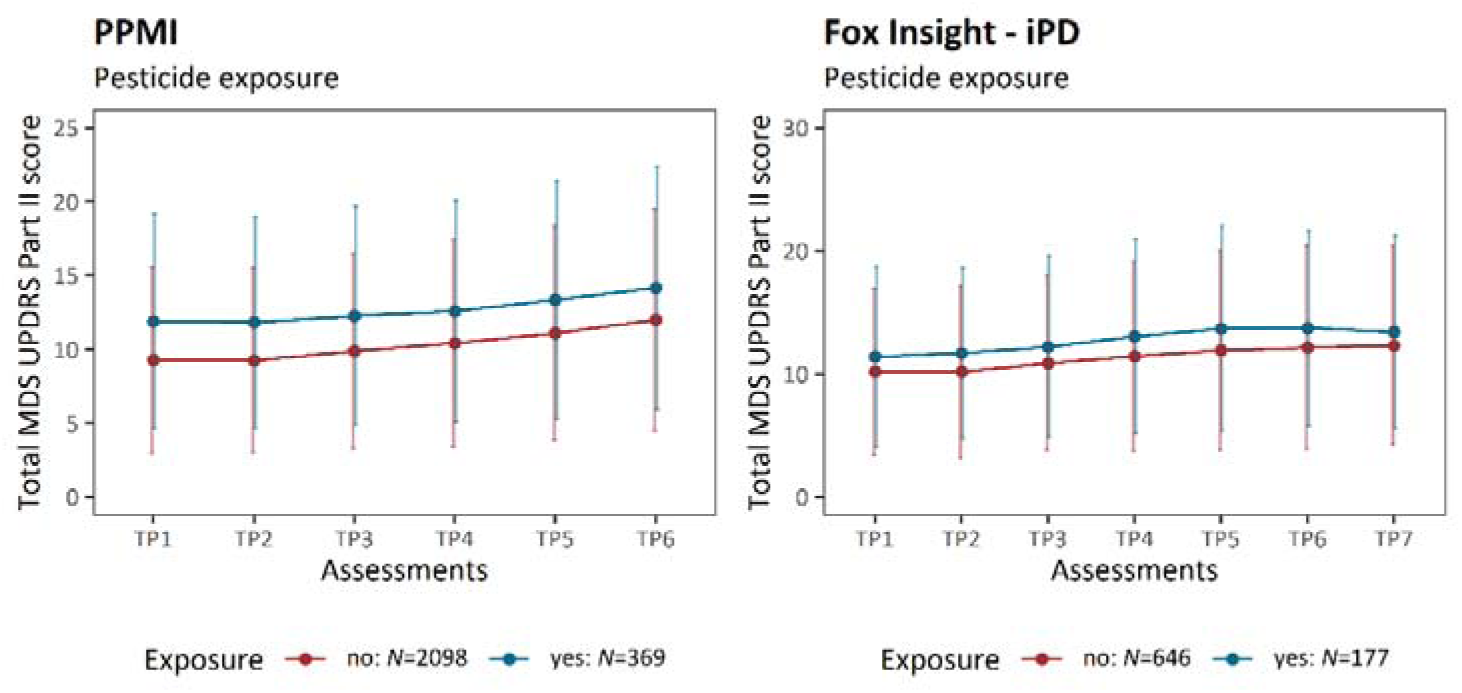
Motor sign severity over time stratified by pesticide exposure. The plots show the progression of PD motor features along the longitudinal assessments. The mean cumulative MDS-UPDRS Part II score is indicated at each time period, and the error bars show the corresponding standard deviation. Patients with iPD are shown (PPMI and Fox Insight). The patients are stratified by pesticide exposure. iPD=idiopathic Parkinson’s disease, TP=Time period, *N*=number of individuals.

**Table 3.**
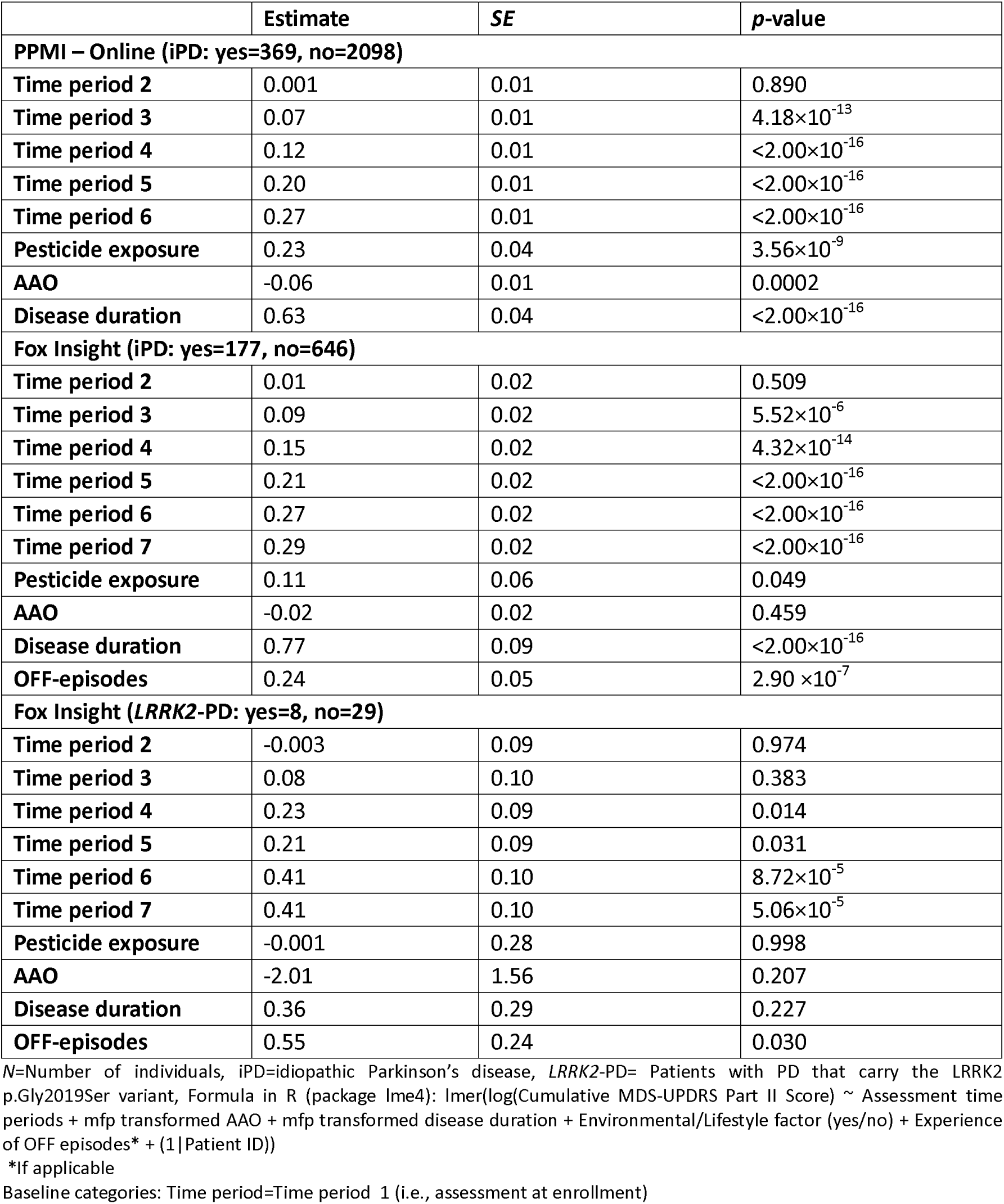
The association between pesticide exposure and motor sign severity over time. The motor sign severity was evaluated longitudinally over time and assessed with a linear mixed model in the PPMI and Fox Insight cohort.

|  | Estimate | SE | p-value |
| --- | --- | --- | --- |
| **PPMI – Online (iPD: yes=369, no=2098)** |  |  |  |
| Time period 2 | 0.001 | 0.01 | 0.890 |
| Time period 3 | 0.07 | 0.01 | $4.18 \times 10^{-13}$ |
| Time period 4 | 0.12 | 0.01 | $<2.00 \times 10^{-16}$ |
| Time period 5 | 0.20 | 0.01 | $<2.00 \times 10^{-16}$ |
| Time period 6 | 0.27 | 0.01 | $<2.00 \times 10^{-16}$ |
| Pesticide exposure | 0.23 | 0.04 | $3.56 \times 10^{-9}$ |
| AAO | -0.06 | 0.01 | 0.0002 |
| Disease duration | 0.63 | 0.04 | $<2.00 \times 10^{-16}$ |
| **Fox Insight (iPD: yes=177, no=646)** |  |  |  |
| Time period 2 | 0.01 | 0.02 | 0.509 |
| Time period 3 | 0.09 | 0.02 | $5.52 \times 10^{-6}$ |
| Time period 4 | 0.15 | 0.02 | $4.32 \times 10^{-14}$ |
| Time period 5 | 0.21 | 0.02 | $<2.00 \times 10^{-16}$ |
| Time period 6 | 0.27 | 0.02 | $<2.00 \times 10^{-16}$ |
| Time period 7 | 0.29 | 0.02 | $<2.00 \times 10^{-16}$ |
| Pesticide exposure | 0.11 | 0.06 | 0.049 |
| AAO | -0.02 | 0.02 | 0.459 |
| Disease duration | 0.77 | 0.09 | $<2.00 \times 10^{-16}$ |
| OFF-episodes | 0.24 | 0.05 | $2.90 \times 10^{-7}$ |
| **Fox Insight (LRRK2-PD: yes=8, no=29)** |  |  |  |
| Time period 2 | -0.003 | 0.09 | 0.974 |
| Time period 3 | 0.08 | 0.10 | 0.383 |
| Time period 4 | 0.23 | 0.09 | 0.014 |
| Time period 5 | 0.21 | 0.09 | 0.031 |
| Time period 6 | 0.41 | 0.10 | $8.72 \times 10^{-5}$ |
| Time period 7 | 0.41 | 0.10 | $5.06 \times 10^{-5}$ |
| Pesticide exposure | -0.001 | 0.28 | 0.998 |
| AAO | -2.01 | 1.56 | 0.207 |
| Disease duration | 0.36 | 0.29 | 0.227 |
| OFF-episodes | 0.55 | 0.24 | 0.030 |

N=Number of individuals, iPD=idiopathic Parkinson's disease, LRRK2-PD= Patients with PD that carry the LRRK2 p.Gly2019Ser variant, Formula in R (package lme4): lmer(log(Cumulative MDS-UPDRS Part II Score) ~ Assessment time periods + mfp transformed AAO + mfp transformed disease duration + Environmental/Lifestyle factor (yes/no) + Experience of OFF episodes\* + (1| Patient ID))
\*if applicable
Baseline categories: Time period=Time period 1 (i.e., assessment at enrollment)

In accordance with the data from the PPMI Online cohort, we observed that iPD patients had visibly more severe motor signs when they were exposed to pesticides (mean cumulative MDS-UPDRS Part II (±*SD*)=13.42 (±7.84)) compared to those that were not (mean cumulative MDS-UPDRS Part II (±*SD*)=12.33 (±8.04), **Figure 3**). The association between pesticide exposure and increased motor sign severity over time in iPD patients from Fox Insight was not significant after adjusting for multiple testing but the trend was in the same direction (β=0.11, *SE*=0.06, *p*=0.049, **Table 3**). It is important to note that the linear mixed effects model was adjusted based on the experience of OFF episodes in the Fox Insight cohort but not in the PPMI Online data, as this information was unavailable. The prevalence of pesticide exposure in a work setting was higher among iPD patients from Fox Insight (∼22%, *N*=823) compared to PPMI Online.

In contrast, we did not observe an association between pesticide exposure and disease severity in patients with *LRRK2*-PD (β=-0.001, *SE*=0.28, *p*=0.998, *N*=37).

### 3.3 Smoking Behavior

The prevalence of smoking was ∼35% among the patients with iPD in the PPMI Online cohort (*N*=2576). Interestingly, smoking status was associated with increased disease severity longitudinally (β=0.13, *SE*=0.03, *p*=9.65×10^-6^) (**Table 4**). At the assessment time period six (i.e., 27-35 months), patients who smoked had more severe motor signs (mean cumulative MDS-UPDRS Part II (±*SD*)=13.14 (±7.71)) compared to those who did not (mean cumulative MDS-UPDRS Part II (±*SD*)=11.64 (±7.48)) (**Figure 4**).

**Figure 4.**
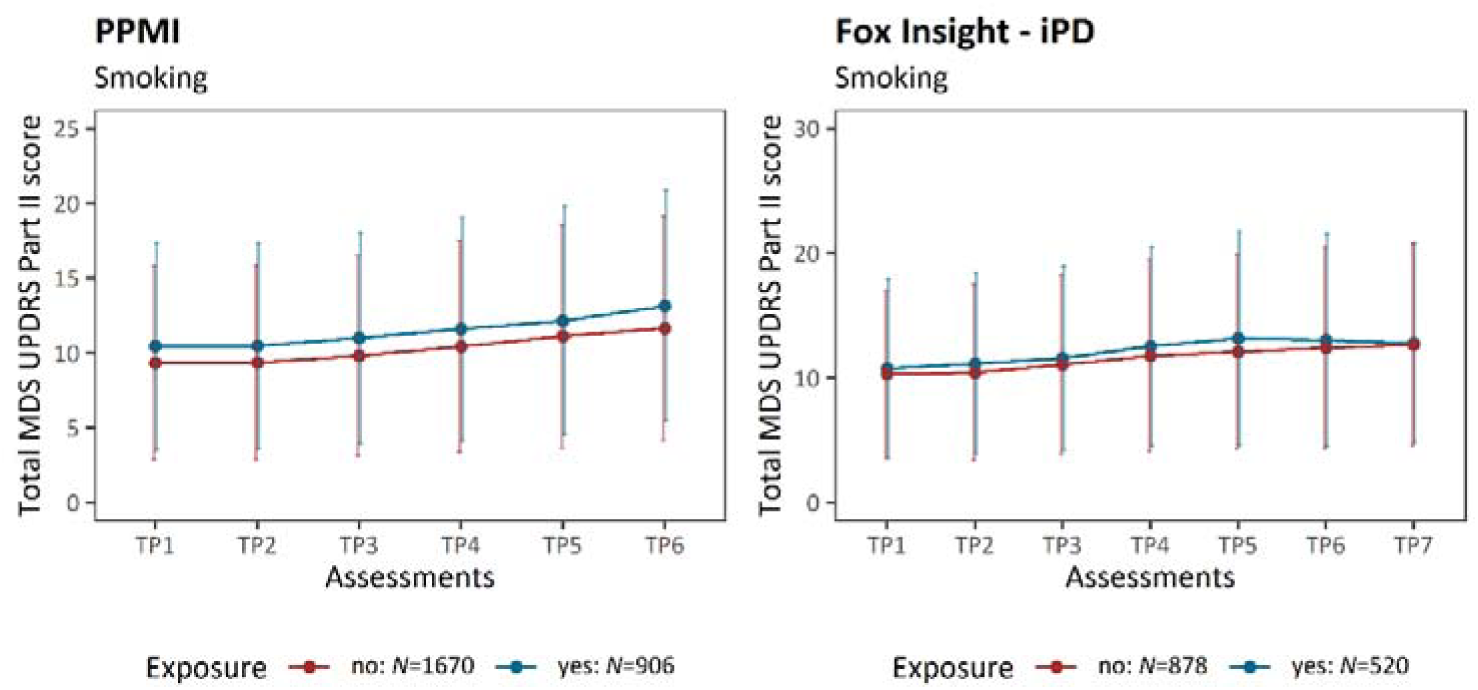
Motor sign severity over time stratified by smoking. The plots show the progression of PD motor features along the longitudinal assessments. The mean cumulative MDS-UPDRS Part II score is indicated at each time period, and the error bars show the corresponding standard deviation. Patients with iPD are shown (PPMI and Fox Insight). The patients are stratified by smoking status. iPD=idiopathic Parkinson’s disease, TP=Time period, *N*=number of individuals.

**Table 4.** The association between smoking and motor sign severity over time. The motor sign severity was evaluated longitudinally over time and assessed with a linear mixed model in the PPMI and Fox Insight cohort.

|  | Estimate | SE | p-value |
| --- | --- | --- | --- |
| **PPMI – Online (iPD: yes=906, no=1670)** |  |  |  |
| Time period 2 | 0.001 | 0.01 | 0.911 |
| Time period 3 | 0.07 | 0.01 | $1.95 \times 10^{-11}$ |
| Time period 4 | 0.12 | 0.01 | $<2.00 \times 10^{-16}$ |
| Time period 5 | 0.20 | 0.01 | $<2.00 \times 10^{-16}$ |
| Time period 6 | 0.26 | 0.01 | $<2.00 \times 10^{-16}$ |
| Smoking | 0.13 | 0.03 | $9.65 \times 10^{-6}$ |
| AAO | -0.05 | 0.01 | 0.001 |
| Disease duration | 0.63 | 0.04 | $<2.00 \times 10^{-16}$ |
| **Fox Insight (iPD: yes=520, no=878)** |  |  |  |
| Time period 2 | 0.02 | 0.01 | 0.258 |
| Time period 3 | 0.08 | 0.01 | $5.77 \times 10^{-9}$ |
| Time period 4 | 0.16 | 0.01 | $<2.00 \times 10^{-16}$ |
| Time period 5 | 0.23 | 0.02 | $<2.00 \times 10^{-16}$ |
| Time period 6 | 0.28 | 0.02 | $<2.00 \times 10^{-16}$ |
| Time period 7 | 0.31 | 0.02 | $<2.00 \times 10^{-16}$ |
| Smoking | 0.06 | 0.04 | 0.122 |
| AAO | -0.03 | 0.02 | 0.118 |
| Disease duration | 0.76 | 0.07 | $<2.00 \times 10^{-16}$ |
| OFF-episodes | 0.24 | 0.04 | $4.19 \times 10^{-11}$ |
| **Fox Insight (LRRK2-PD: yes=22, no=28)** |  |  |  |
| Time period 2 | -0.02 | 0.07 | 0.831 |
| Time period 3 | 0.05 | 0.08 | 0.493 |
| Time period 4 | 0.20 | 0.08 | 0.008 |
| Time period 5 | 0.16 | 0.08 | 0.035 |
| Time period 6 | 0.34 | 0.08 | $5.30 \times 10^{-5}$ |
| Time period 7 | 0.37 | 0.08 | $1.02 \times 10^{-5}$ |
| Smoking | -0.07 | 0.20 | 0.735 |
| AAO | -0.88 | 1.26 | 0.488 |
| Disease duration | 0.35 | 0.25 | 0.165 |
| OFF-episodes | 0.57 | 0.21 | 0.009 |

N=Number of individuals, iPD=idiopathic Parkinson's disease, LRRK2-PD= Patients with PD that carry the LRRK2 p.Gly2019Ser variant, Formula in R (package lme4): lmer(log(Cumulative MDS-UPDRS Part II Score) ~ Assessment time periods + mfp transformed AAO + mfp transformed disease duration + Environmental/Lifestyle factor (yes/no) + Experience of OFF episodes\* + (1|Patient ID))
\*If applicable
Baseline categories: Time period=Time period 1 (i.e., assessment at enrollment)

We observed a subtle trend for increased motor sign severity over time in smokers from the Fox Insight cohort, and this trend was limited to patients with iPD (β=0.06, *SE*=0.04, *p*=0.122, *N*=1398) and not present in *LRRK2*-PD (β=-0.07, *SE*=0.20, *p*=0.735, *N*=50) (**Table 4**).

### 3.4 Caffeinated Beverage Consumption

Next, we investigated the longitudinal correlation between four caffeinated beverages (i.e., black tea, caffeinated soda, coffee, and green tea) and motor sign severity. Black tea consumption was similar among the iPD patients of the PPMI Online and Fox Insight cohort (PPMI Online: ∼40%, *N*=2555; Fox Insight: 39%, *N*=1213). However, we observed no difference in the disease severity over time when stratifying for black tea consumption (**Figure 5**, **Table 5**).

**Figure 5.**
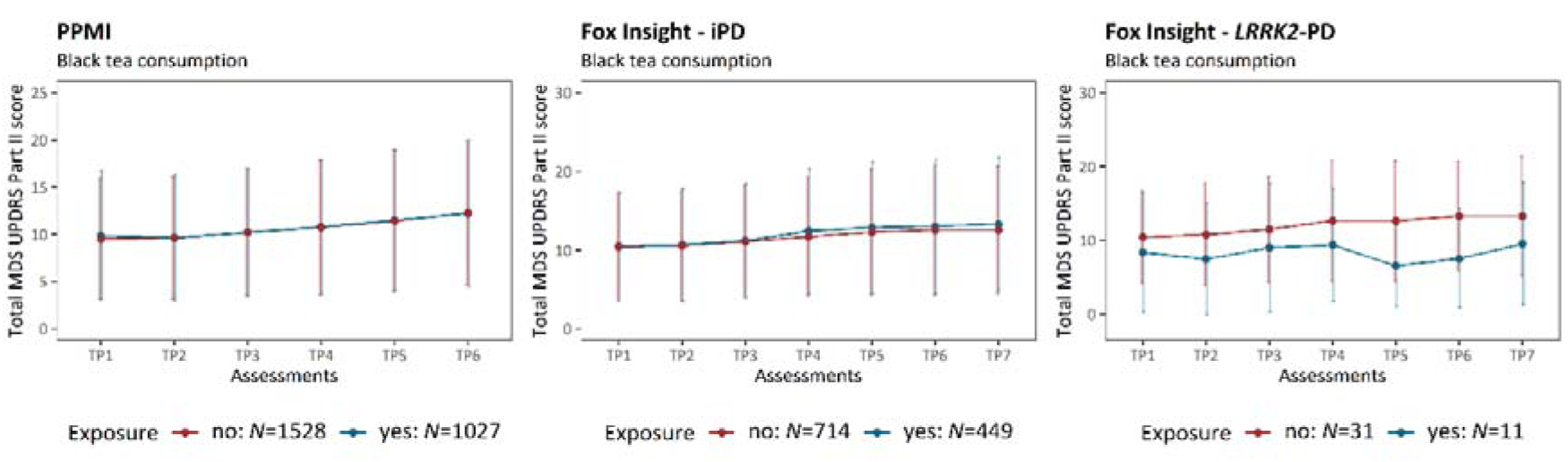
Motor sign severity over time stratified by black tea consumption. The plots show the progression of PD motor features along the longitudinal assessments. The mean cumulative MDS-UPDRS Part II score is indicated at each time period, and the error bars show the corresponding standard deviation. Patients with iPD (PPMI and Fox Insight) and *LRRK2*-PD (Fox Insight) are shown. The patients are stratified by black tea consumption. iPD=idiopathic Parkinson’s disease, *LRRK2*-PD=Patients with PD that carry the LRRK2 p.Gly2019Ser variant, TP=Time period, *N*=number of individuals.

**Table 5.**
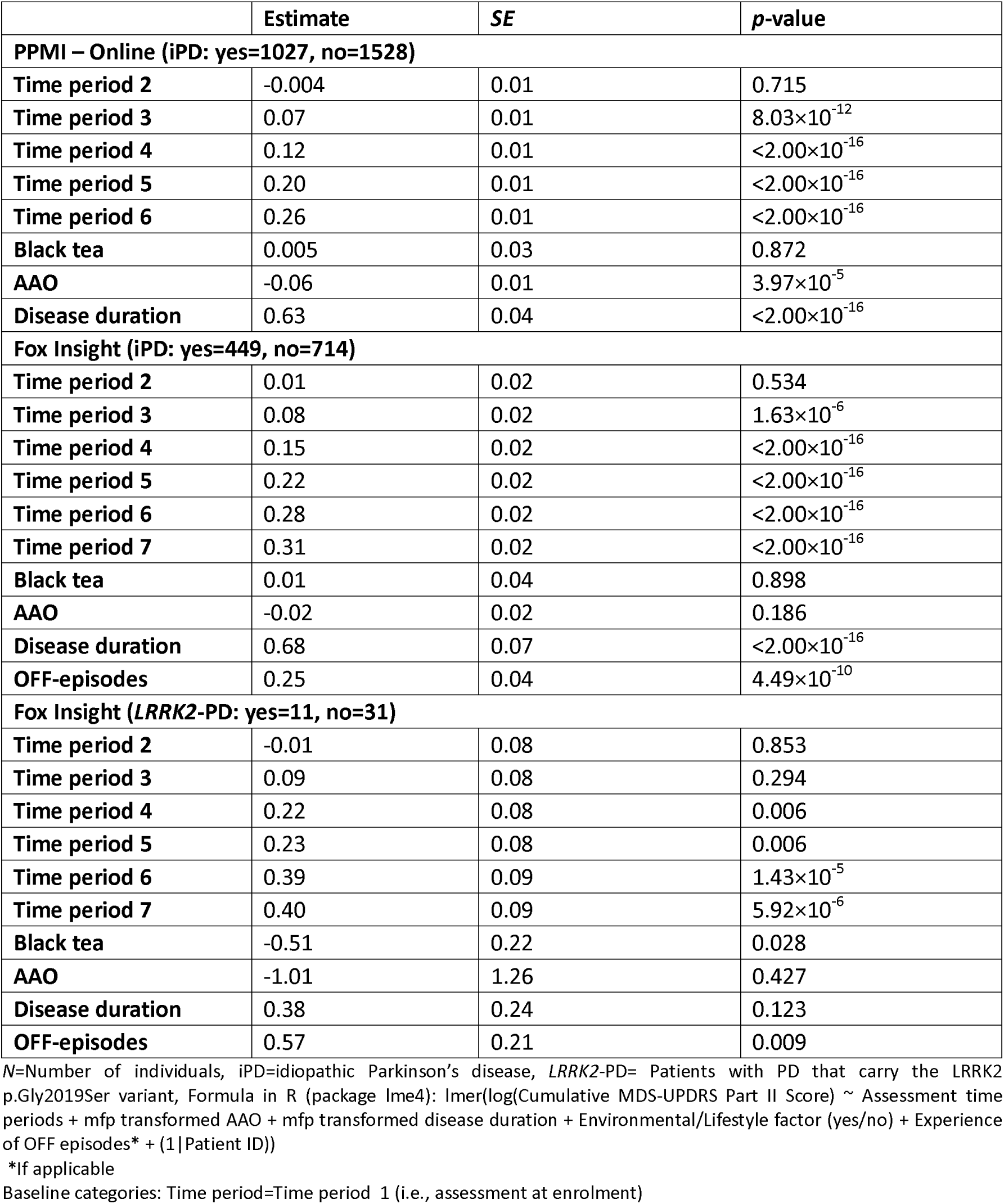
The association between black tea consumption and motor sign severity over time. The motor sign severity was evaluated longitudinally over time and assessed with a linear mixed model in the PPMI and Fox Insight cohort.

Interestingly, we specifically observed a large difference in motor sign severity after 60-month enrollment between *LRRK2*-PD patients who consumed black tea (mean cumulative MDS-UPDRS Part II (±*SD*)=9.50 (±8.32)) and those who did not (mean cumulative MDS-UPDRS Part II (±*SD*)=13.27 (±8.11)). The linear mixed effects model confirmed the association between black tea and less severe motor signs (β=-0.51, *SE*=0.22, *p*=0.028) (**Table 5**). Although the sample size of *LRRK2*-PD patients with available environment and lifestyle data was small (*N*=42, prevalence of black tea consumption: ∼26%), the effect size was the largest out of all included environment/lifestyle factors. Additionally, when including all iPD and *LRRK2*-PD patients from Fox Insight into one model and assessing the joint effect on disease severity of *LRRK2*-PD/iPD status and black tea consumption (i.e., LRRK2-PD/iPD status × Black tea consumption), there was evidence for an interaction effect between PD type (iPD or *LRRK2*-PD) and black tea consumption (β=-0.51, *SE*=0.26, *p*=0.045, *N*=1333).

In contrast to black tea consumption, caffeinated soda was associated with more severe motor signs. The prevalence of caffeinated soda consumption was ∼55% (*N*=2559) in iPD patients of PPMI Online. After the 35-month enrollment time, iPD patients who did drink caffeinated soda had more severe motor signs (mean cumulative MDS-UPDRS Part II (±*SD*)=12.85 (±7.93)) compared to those who did not (mean cumulative MDS-UPDRS Part II (±*SD*)=11.47 (±7.37)) and the association was confirmed by the linear mixed model (β=0.15, *SE*=0.03, *p*=3.84×10^-8^) (**Figure 6**, **Table 6**).

**Figure 6.**
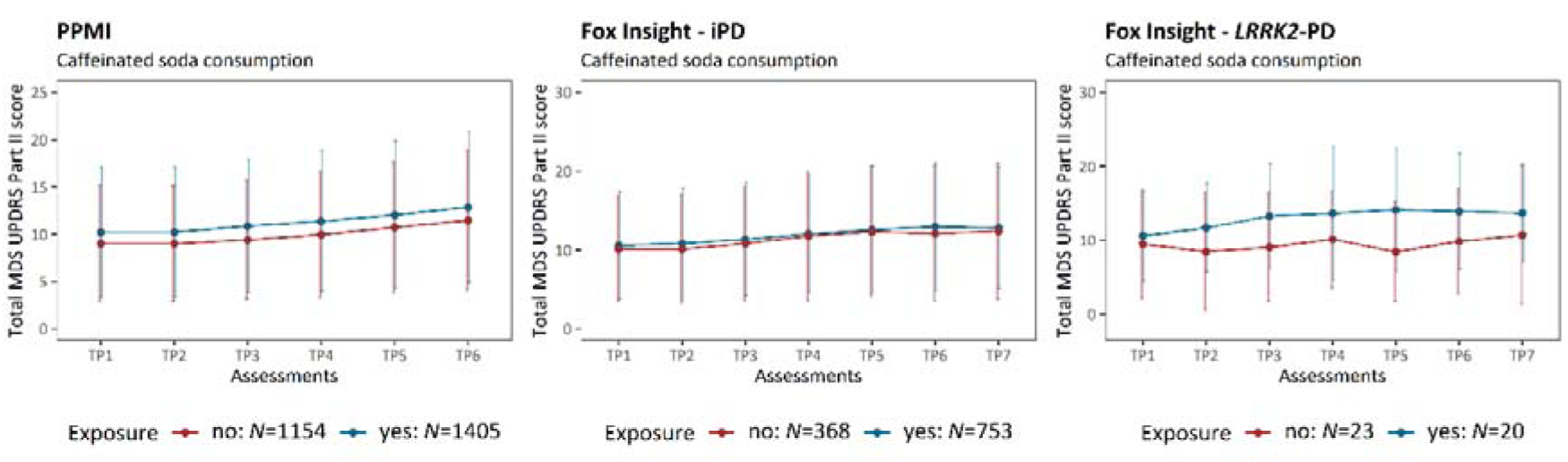
Motor sign severity over time stratified by caffeinated soda consumption. The plots show the progression of PD motor features along the longitudinal assessments. The mean cumulative MDS-UPDRS Part II score is indicated at each time period, and the error bars show the corresponding standard deviation. Patients with iPD (PPMI and Fox Insight) and *LRRK2*-PD (Fox Insight) are shown. The patients are stratified by caffeinated soda consumption. iPD=idiopathic Parkinson’s disease, *LRRK2*-PD=Patients with PD that carry the LRRK2 p.Gly2019Ser variant, TP=Time period, *N*=number of individuals.

**Table 6.** The association between caffeinated soda consumption and motor sign severity over time. The motor sign severity was evaluated longitudinally over time and assessed with a linear mixed model in the PPMI and Fox Insight cohort.

|  | Estimate | SE | p-value |
| --- | --- | --- | --- |
| **PPMI – Online (iPD: yes=1405, no=1154)** |  |  |  |
| Time period 2 | 0.0005 | 0.01 | 0.964 |
| Time period 3 | 0.07 | 0.01 | $1.18 \times 10^{-12}$ |
| Time period 4 | 0.12 | 0.01 | $<2.00 \times 10^{-16}$ |
| Time period 5 | 0.20 | 0.01 | $<2.00 \times 10^{-16}$ |
| Time period 6 | 0.26 | 0.01 | $<2.00 \times 10^{-16}$ |
| Caffeinated soda | 0.15 | 0.03 | $3.84 \times 10^{-8}$ |
| AAO | -0.06 | 0.01 | $3.04 \times 10^{-5}$ |
| Disease duration | 0.63 | 0.04 | $<2.00 \times 10^{-16}$ |
| **Fox Insight (iPD: yes=753, no=368)** |  |  |  |
| Time period 2 | 0.01 | 0.02 | 0.533 |
| Time period 3 | 0.08 | 0.02 | $5.48 \times 10^{-7}$ |
| Time period 4 | 0.15 | 0.02 | $<2.00 \times 10^{-16}$ |
| Time period 5 | 0.22 | 0.02 | $<2.00 \times 10^{-16}$ |
| Time period 6 | 0.28 | 0.02 | $<2.00 \times 10^{-16}$ |
| Time period 7 | 0.31 | 0.02 | $<2.00 \times 10^{-16}$ |
| Caffeinated soda | 0.09 | 0.04 | 0.031 |
| AAO | -0.02 | 0.02 | 0.189 |
| Disease duration | 0.72 | 0.07 | $<2.00 \times 10^{-16}$ |
| OFF-episodes | 0.24 | 0.04 | $2.22 \times 10^{-9}$ |
| **Fox Insight (LRRK2-PD: yes=20, no=23)** |  |  |  |
| Time period 2 | -0.01 | 0.08 | 0.882 |
| Time period 3 | 0.09 | 0.08 | 0.260 |
| Time period 4 | 0.22 | 0.08 | 0.006 |
| Time period 5 | 0.23 | 0.08 | 0.005 |
| Time period 6 | 0.39 | 0.09 | $1.18 \times 10^{-5}$ |
| Time period 7 | 0.40 | 0.08 | $4.98 \times 10^{-6}$ |
| Caffeinated soda | 0.35 | 0.21 | 0.108 |
| AAO | -1.07 | 1.31 | 0.419 |
| Disease duration | 0.46 | 0.24 | 0.069 |
| OFF-episodes | 0.43 | 0.23 | 0.070 |

N=Number of individuals, iPD=idiopathic Parkinson's disease, LRRK2-PD= Patients with PD that carry the LRRK2 p.Gly2019Ser variant, Formula in R (package lme4): lmer(log(Cumulative MDS-UPDRS Part II Score) ~ Assessment time periods + mfp transformed AAO + mfp transformed disease duration + Environmental/Lifestyle factor (yes/no) + Experience of OFF episodes\* + (1|Patient ID))
\*If applicable
Baseline categories: Time period=Time period 1 (i.e., assessment at enrolment)

In patients with iPD from the Fox Insight cohort, the prevalence of caffeinated soda consumption was ∼67% (*N*=1121). After an enrollment time of 60 months, the iPD patients from Fox Insight who consumed caffeinated soda had more severe motor signs (mean cumulative MDS-UPDRS Part II (±*SD*)=12.79 (±7.72)) compared to those who did not as well (mean cumulative MDS-UPDRS Part II (±*SD*)=12.41 (±8.70)) and the association was confirmed by the linear mixed model (β=0.09, *SE*=0.04, *p*=0.031). With regard to the *LRRK2*-PD patients (prevalence of caffeinated soda consumption: ∼47%, *N*=43), patients who consumed caffeinated soda had visibly more severe motor signs (mean cumulative MDS-UPDRS Part II (±*SD*)=13.67 (±6.61)) compared to the *LRRK2*-PD patients that did not (mean cumulative MDS-UPDRS Part II (±*SD*)=10.71 (±9.43)) (**Figure 6**). However, the association was not confirmed when the linear mixed model was applied (β=0.35, *SE*=0.21, *p*=0.108) (**Table 6**).

Coffee and green tea consumption were explored analogously to the other caffeinated beverages; however, we did not observe an association with disease severity over time in patients with iPD or *LRRK2*-PD (**Supplementary Table 1-2**).

## 4. Discussion

Investigating the impact of environmental and lifestyle factors beyond the onset of PD is of great importance, as there is increasing evidence that those factors affect the disease course. Thus far, there has been no longitudinal study on the association between smoking, caffeine consumption, and PD features severity. Therefore, we utilized large available data sets with longitudinal follow-ups of patients with PD (i.e., PPMI-Online and Fox Insight) to thoroughly investigate the effect of pesticide exposure, smoking and caffeine over time. Additionally, we explored the association between environment, lifestyle and disease severity in patients with monogenic *LRRK2*-PD in comparison to iPD. Longitudinal cohorts have major advantages over studies with only one study time point, as exposures are recorded at an early study stage (e.g., Fox Insight at study enrollment), resulting in a higher potential of a causal relationship. In the case of PPMI-Online, the assessment of environmental/lifestyle factors was not done at the study enrollment but at different time points throughout the study. However, we utilized the assessment of PPMI-Online, in which the largest number of participants answered the respective environmental and lifestyle questionnaires, to maximize the sample size and statistical power.

The progression in the MDS-UPDRS part II score during the longitudinal assessment was in the expected range (Holden et al., 2018), but patients with *LRRK2*-PD had less severe motor signs than patients with iPD. This finding concords with a recent 3.5-year longitudinal study demonstrating less severe PD features in patients with *LRRK2*-PD (Kmiecik et al., 2024). They found that patients with iPD and *LRRK2*-PD differed the most in their non-motor PD features, and *LRRK2*-PD patients presented with predominantly motor impairment. Here, we could show that patients with *LRRK2*-PD consistently present with less severe motor features over the complete enrollment time as well (**Figure 2**).

### 4.1 Environmental Exposures and Lifestyle Factors are Longitudinally Associated with Disease Severity

Pesticide exposure is the most substantial environmental risk factor for PD (Noyce et al., 2012) and is associated with an earlier disease onset as well (Gamache et al., 2019; Ratner et al., 2014). In line with increased risk and earlier AAO, we found that pesticide exposure was significantly associated with increased motor signs in iPD, thus further underlying the link between pesticides and PD pathogenesis. Our study adds to the increasing evidence that pesticide exposure impacts patients with PD beyond the onset of the disease, as another longitudinal study recently found specific pesticides to be associated with faster PD progression (Li et al., 2023).

Our study demonstrated the association between occupational pesticide exposure and motor signs in the large PPMI-Online cohort. Although the association between exposure and more severe motor signs did not reach significance in the Fox Insight cohort, the trend was in the same direction and iPD patients that were exposed to pesticides had a visible higher motor sign severity over time as well. However, we did not detect an association in *LRRK2*-PD, thus they might not be as vulnerable to pesticide exposure. Although our sample size of *LRRK2*-PD patients with available pesticide information was limited and therefore, the statistical power was low, recently, a larger-sized study found no association between LRRK2 G2019S penetrance and pesticide exposure, rather an increased disease risk among carriers of PD-related *GBA1* variants (Brown et al., 2024). Classically, pesticides like rotenone and paraquat are known to impair mitochondrial function (Grunewald et al., 2019), and there is increasing evidence that lysosomal and autophagy dysfunction has been shown to increase PD susceptibility in persons with heavy pesticide exposure (Ngo et al., 2024). It is important to note that lysosomal autophagic dysfunction is one of the key players in the pathogenesis of *GBA1*-related PD (Kim et al., 2018; Pitcairn et al., 2019; Pradas and Martinez-Vicente, 2023), which might explain the increased PD risk in *GBA1*. Although lysosomal impairment has also been observed in *LRRK2*-related PD (Bonet-Ponce and Cookson, 2022; Schapansky et al., 2014; Yadavalli and Ferguson, 2023), the pathogenesis of *LRRK2*-PD is much more diverse and less well elucidated including mitochondrial dysfunction, vesicle trafficking and protein scaffolding among other pathways (Delcambre et al., 2020; Qi et al., 2023; Taymans et al., 2015).

The harmful impact of pesticide exposure, even beyond the onset of PD, is particularly important to highlight. In contrast to unfavorable lifestyle factors, which can be optimized in patients or persons at risk, occupational or residential pesticide exposure is much harder to avoid, and exposure can happen unconsciously. In our longitudinal study, we focused on occupational pesticide exposure, as the data was available in both assessed longitudinal cohorts. Occupational exposure is less prevalent in the population than residential exposure. Additionally, recall biases might impact self-reported exposure, which applies to all self-reported environmental or lifestyle data.

Smoking is one of the lifestyle factors with the most considerable protective effect on PD (Noyce et al., 2012), and is associated with a later AAO in iPD and *LRRK2*-PD (Gabbert et al., 2022; Gigante et al., 2017; Luth et al., 2023), and the protective effect might even be causal (Grover et al., 2019). In contrast, smoking was associated with more severe motor signs over time in our study. The association was detected in iPD patients from PPMI-Online, but there was also a visible trend for increased disease severity in iPD patients from Fox Insight. Compared to pesticide exposure, the effect size was smaller and the association was only present in one of the cohorts. Therefore, smoking might not be as robustly associated with more severe motor signs. Nevertheless, the results of our longitudinal study support the findings from a cross-sectional assessment of smoking and increased disease severity (Gabbert et al., 2023).

Our study presents evidence that the protective effect of smoking might be restricted to the time window before the disease onset. Indeed, smoking is associated with increased all-cause mortality and various health consequences (Gellert et al., 2012; Warren et al., 2014). Therefore, it may be the case that the overall harmfulness of smoking overshadows the PD-specific protective effects after disease onset. Interestingly, it has recently been shown that the time window for the beneficial neuroprotective effects of nicotine is very small and likely ineffective after the majority of dopaminergic neurons are lost (Deliz et al., 2024; Mannett et al., 2022).

Lastly, we assessed caffeine consumption, including coffee, black tea, green tea, and caffeinated soda. Coffee and tea are also considered protective lifestyle factors in the context of PD (Noyce et al., 2012) and our group previously reported a strong correlation between black tea consumption and later AAO in patients with *LRRK2*-PD (Luth et al., 2020). We did not observe any association between coffee or green tea and disease severity over time in our study cohorts. However, black tea was associated explicitly with less severe motor signs in *LRRK2*-PD, and there was no difference in patients with iPD stratified for black tea consumption. As the sample size of longitudinally followed-up patients with *LRRK2*-PD and available lifestyle data in the Fox Insight cohort was limited, we investigated the association between motor signs and black tea consumption in an independent longitudinal cohort. Therefore, we utilized the PPMI cohort (Marek et al., 2018) and only participants not enrolled in the PPMI-Online sub-study (*LRRK2*-PD: *N*= 39, enrollment time=90 months). Interestingly, we could confirm the association between black tea consumption and less severe motor signs in patients with *LRRK2*-PD (β=-0.41, *SE*=0.18, *p*=0.026) (**Supplementary Figure 4**, **Supplementary Table 3**). It is important to note that black tea consumption was the only factor in our study associated with decreased disease severity over time, and the association was robustly present in two independent cohorts. Subsequently, the protective effect of black tea consumption might remain present after disease onset. The underlying molecular pathway of the potential protective effect of black tea consumption is not fully understood. There is evidence that specific tea components have a neuroprotective effect (Mu et al., 2021) and prevent apoptosis of dopaminergic neurons by increasing mitochondrial biogenesis (Jhuo et al., 2024). Indeed, mitochondrial dysfunction is essential for PD pathogenesis; still, it might not affect all patients with PD. Thus, black tea consumption potentially has a larger effect on *LRRK2*-PD patients compared to the inhomogeneous group of patients with iPD.

In contrast to black tea, we found that caffeinated soda consumption was consistently associated with more severe motor signs in patients with iPD from PPMI Online and Fox Insight, underscoring the association’s robustness. In addition, there was also a strong visible trend for more severe motor signs in *LRRK2*-PD patients who consumed caffeinated soda, but the association did not reach significance. Our group has previously shown that caffeinated soda, different from other caffeinated beverages, is not a protective factor in PD and is associated with earlier disease onset in patients with *LRRK2*-PD (Luth et al., 2020) and might increase the vulnerability of *LRRK2*-PD patients for increased mitochondrial dysfunctions (Luth et al., 2023). Thus far, the underlying molecular pathway mediating the effects of caffeinated soda is unknown. However, as coffee and tea are protective lifestyle factors in PD, caffeine-independent mechanisms might be involved.

Comparing black tea and caffeinated sodas and their opposite effect on disease severity over time, sodas probably differ from tea in terms of ingredients and higher sugar content. In the last years, type 2 diabetes (T2DM) has been elucidated as an important comorbidity in the context of PD (Chohan et al., 2021). T2DM has even been associated with more severe motor signs (Athauda et al., 2022), and GLP1 receptor agonists, which are T2DM treatments, have recently been found to hold the progression of motor signs in PD patients (Meissner et al., 2024). Certainly, particular lifestyle and dietary behaviors can also increase the risk of developing T2DM. Unfortunately, the sample size of patients with PD and T2DM in our study cohorts was too small to explore the joint effects of T2DM and different lifestyle factors. Still, potential interactions should be considered for future studies. The underlying pathway of how caffeinated soda or black tea potentially affects disease severity has not been elucidated yet. However, our study is the first to present evidence that these caffeinated beverages are longitudinally associated with motor sign severity and thus may impact PD patients beyond the disease onset.

### 4.2 Strengths and Limitations

The main strength of our study is the large sample size of patients with iPD who were consistently followed up over a long period of time. Evaluating the association between environment, lifestyle and disease severity over time at consecutive points in time allows a more comprehensive assessment compared to a cross-sectional one. Investigating the environment and lifestyle in two cohorts demonstrates the reproducibility and robustness of our observed associations. The availability of genetic data for two PD subtypes in Fox Insight made it possible to distinguish between iPD and *LRRK2*-PD, which is essential as certain environmental or lifestyle factors may affect patients with other Parkinson’s subtypes differently. Most participants in the study cohorts were patients with iPD, resulting in a small sample size of *LRRK2*-PD patients to explore. Additionally, our study focuses on patients with iPD and *LRRK2*-PD, neglecting other monogenic forms of PD. Another limitation is that recall bias can impact the assessment of environmental and lifestyle factors, as included in this study. In addition, pesticide exposure can even happen without the participants’ knowledge. More objective ways would be utilizing geographical databases of pesticide usage or personal sensor measuring devices, which can benefit an accurate assessment of the environment and lifestyle. Larger studies, including sufficient sample sizes of other monogenic PD forms, will also be required to elucidate and differentiate the impact of environmental factors on different PD subtypes and their disease progression.

### 4.3 Conclusion

The results of our longitudinal study provide further evidence of the importance of environment and lifestyle in PD, even after the disease onset. We observed that pesticide exposure and caffeinated soda are associated with more severe motor signs in iPD. Additionally, smoking might also be associated with disease severity but with a smaller effect size. In patients with *LRRK2*-PD, primarily black tea consumption was associated with less severe motor signs, and the protective effect was specific to *LRRK2*-PD and not present in iPD. Thus, environmental exposure and lifestyle factors affect disease severity in patients with *LRRK2*-PD and iPD. Nevertheless, further replication with larger sample sizes of monogenic PD will be required alongside functional studies elucidating the underlying molecular pathways of how disease severity is impacted by specific environmental exposures or lifestyle factors.

## Data Availability

Data used in the preparation of this article were obtained from the Fox Insight database (https://foxinsight-info.michaeljfox.org/insight/explore/insight.jsp) on 24/07/2024. For up-to-date information on the study, visit https://foxinsight-info.michaeljfox.org/insight/explore/insight.jsp.
Data used in the preparation of this article were obtained 24/07/2024 from the Parkinson's Progression Markers Initiative (PPMI) database (www.ppmi-info.org/access-dataspecimens/download-data), RRID:SCR 006431. For up-to-date information on the study, visit www.ppmi-info.org.

## Acknowledgments

The Fox Insight Study (FI) is funded by The Michael J. Fox Foundation for Parkinson’s Research. We would like to thank the Parkinson’s community and 23andMe research participants and employees for making this research possible.

PPMI – a public-private partnership – is funded by the Michael J. Fox Foundation for Parkinson’s Research and funding partners, including 4D Pharma, Abbvie, AcureX, Allergan, Amathus Therapeutics, Aligning Science Across Parkinson’s, AskBio, Avid Radiopharmaceuticals, BIAL, BioArctic, Biogen, Biohaven, BioLegend, BlueRock Therapeutics, Bristol-Myers Squibb, Calico Labs, Capsida Biotherapeutics, Celgene, Cerevel Therapeutics, Coave Therapeutics, DaCapo Brainscience, Denali, Edmond J. Safra Foundation, Eli Lilly, Gain Therapeutics, GE HealthCare, Genentech, GSK, Golub Capital, Handl Therapeutics, Insitro, Jazz Pharmaceuticals, Johnson & Johnson Innovative Medicine, Lundbeck, Merck, Meso Scale Discovery, Mission Therapeutics, Neurocrine Biosciences, Neuron23, Neuropore, Pfizer, Piramal, Prevail Therapeutics, Roche, Sanofi, Servier, Sun Pharma Advanced Research Company, Takeda, Teva, UCB, Vanqua Bio, Verily, Voyager Therapeutics, the Weston Family Foundation and Yumanity Therapeutics.

## Data access

Data used in the preparation of this article were obtained from the Fox Insight database (https://foxinsight-info.michaeljfox.org/insight/explore/insight.jsp) on 24/07/2024. For up-to-date information on the study, visit https://foxinsight-info.michaeljfox.org/insight/explore/insight.jsp.

Data used in the preparation of this article were obtained 24/07/2024 from the Parkinson’s Progression Markers Initiative (PPMI) database (www.ppmi-info.org/access-dataspecimens/download-data), RRID:SCR 006431. For up-to-date information on the study, visit www.ppmi-info.org

## Author contribution

**Theresa Lüth:** Data curation, Formal analysis, Methodology, Investigation, Software, Validation, Visualization, Writing – original draft; **Amke Caliebe:** Investigation, Methodology, Supervision, Writing – review and editing; **Carolin Gabbert:** Data curation, Investigation, Writing – review and editing; **Sebastian Sendel:** Investigation, Writing – review and editing; **Björn-Hergen Laabs:** Investigation, Writing – review and editing; **Inke R. König:** Conceptualization, Funding acquisition, Investigation, Project administration, Resources, Writing – review and editing; **Christine Klein:** Conceptualization, Funding acquisition, Project administration, Resources, Writing – review and editing; **Joanne Trinh:** Conceptualization, Funding acquisition, Investigation, Project administration, Resources, Supervision, Writing – original draft

## Supplementary Material

**Supplementary Figure 1.**
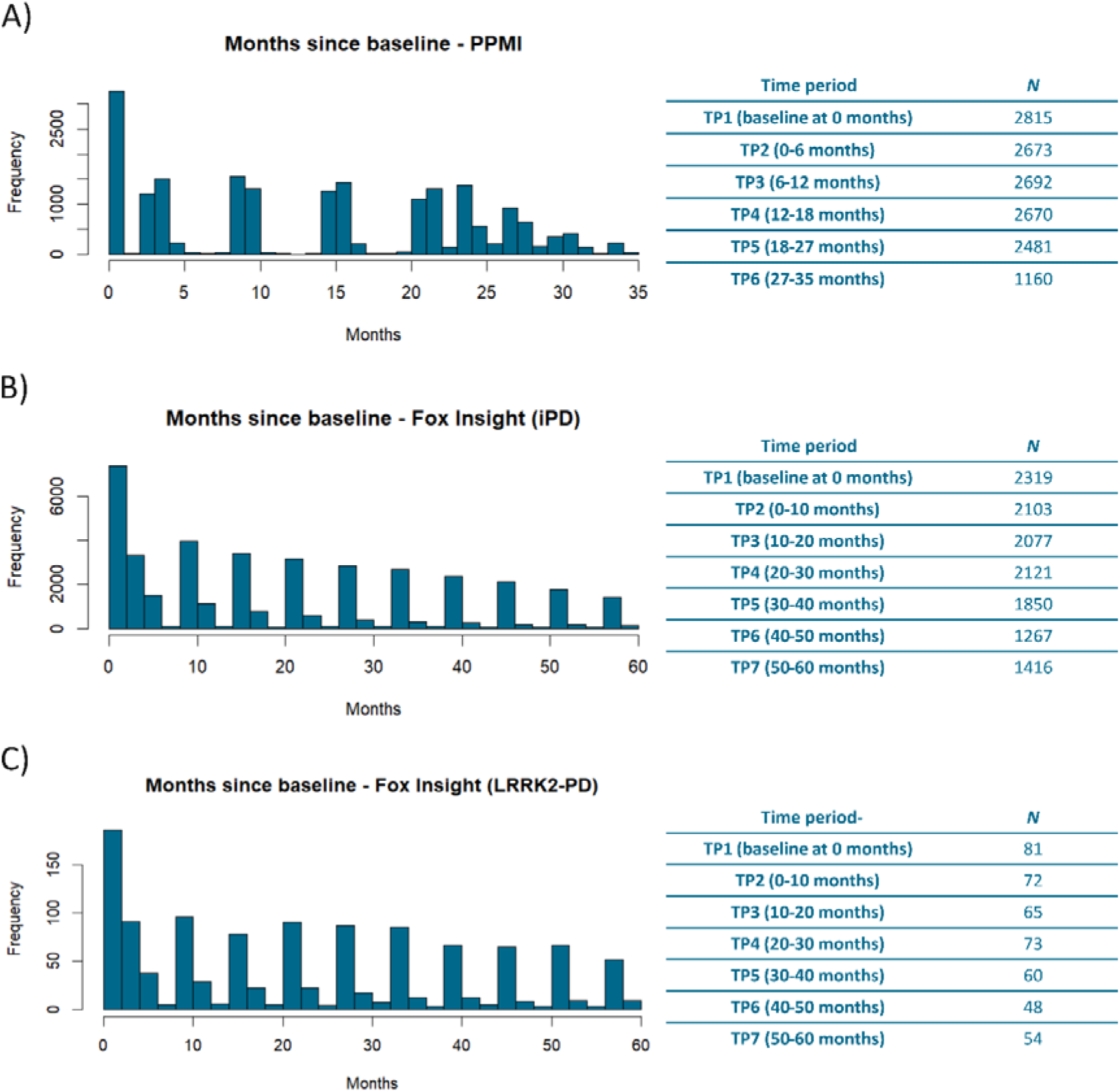
Enrollment of Patients in the PPMI-Online and Fox Insight longitudinal cohort. The bar charts show the number of patients with PD that longitudinally responded to the motor signs assessment in the months since baseline. The assessments were stratified into six time periods (PPMI, **A**) or seven (Fox Insight iPD and LRRK2-PD, **B-C**). The number of patients per assessment is also presented in the corresponding tables. *N*=Number of individuals, iPD=idiopathic Parkinson’s disease, *LRRK2*-PD= Patients with PD that carry the LRRK2 p.Gly2019Ser variant.

**Supplementary Figure 2.**
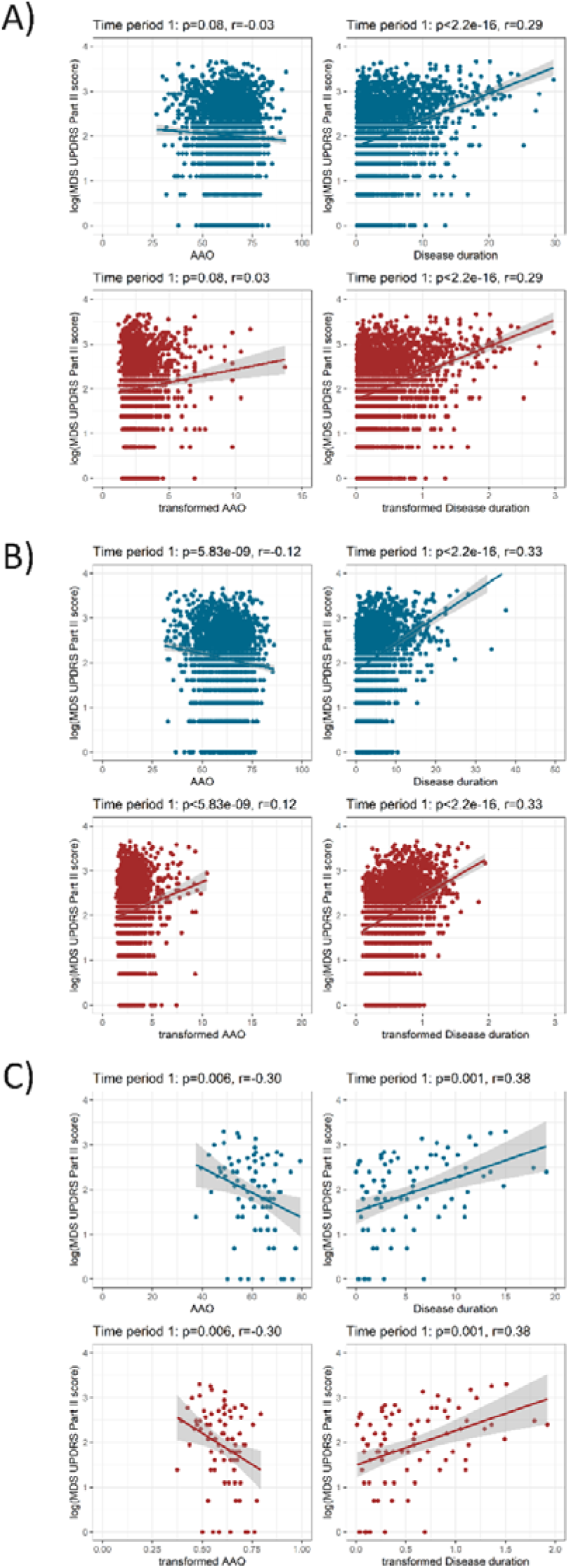
Association between age at onset or disease duration and motor signs score. The scatter plots show the relationship between the age of disease onset (AAO) and disease duration with the logarithmized MDS-UPDRS part II score. The untransformed data are shown in blue, and the transformed data in red (by using fractional polynomials). The plots show the data of patients with iPD (PPMI-Online **A** and Fox Insight in **B**) or with LRRK2-PD (Fox Insight in **C**). iPD=idiopathic Parkinson’s disease, *LRRK2*-PD=Patients with PD that carry the LRRK2 p.Gly2019Ser variant, *r*=Spearman’s rank correlation rho, *p*=Spearman’s rank correlation *p*-value.

**Supplementary Figure 3.**
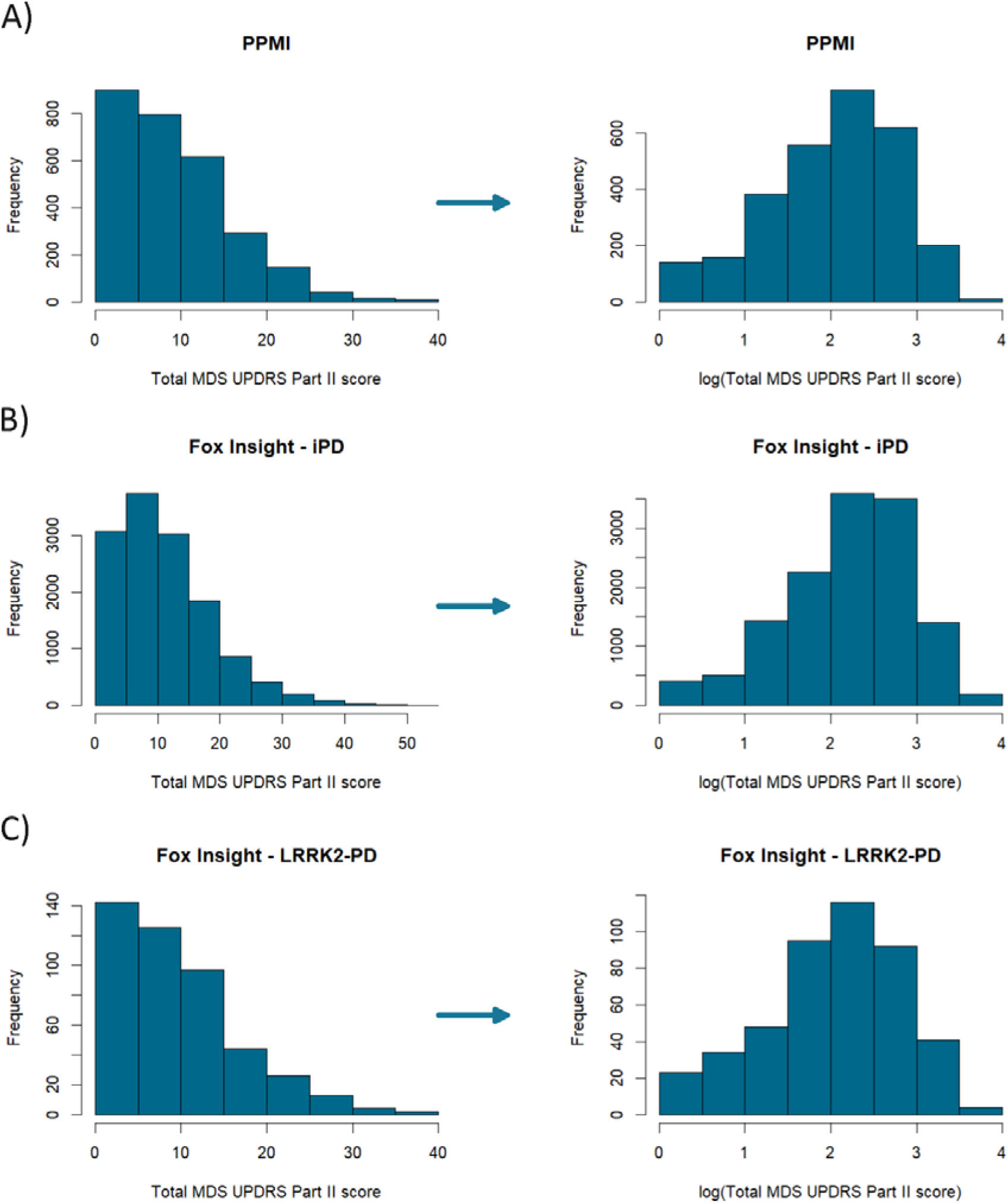
Distribution of the motor signs severity score. The histograms show the frequency of the cumulative MDS-UPDRS Part II score. As there is no normal distribution the score has been logarithmized. The plots show the data of patients with iPD (PPMI-Online **A** and Fox Insight in **B**) or with LRRK2-PD (Fox Insight in **C**). iPD=idiopathic Parkinson’s disease, *LRRK2*-PD=Patients with PD that carry the LRRK2 p.Gly2019Ser variant.

**Supplementary Figure 4.**
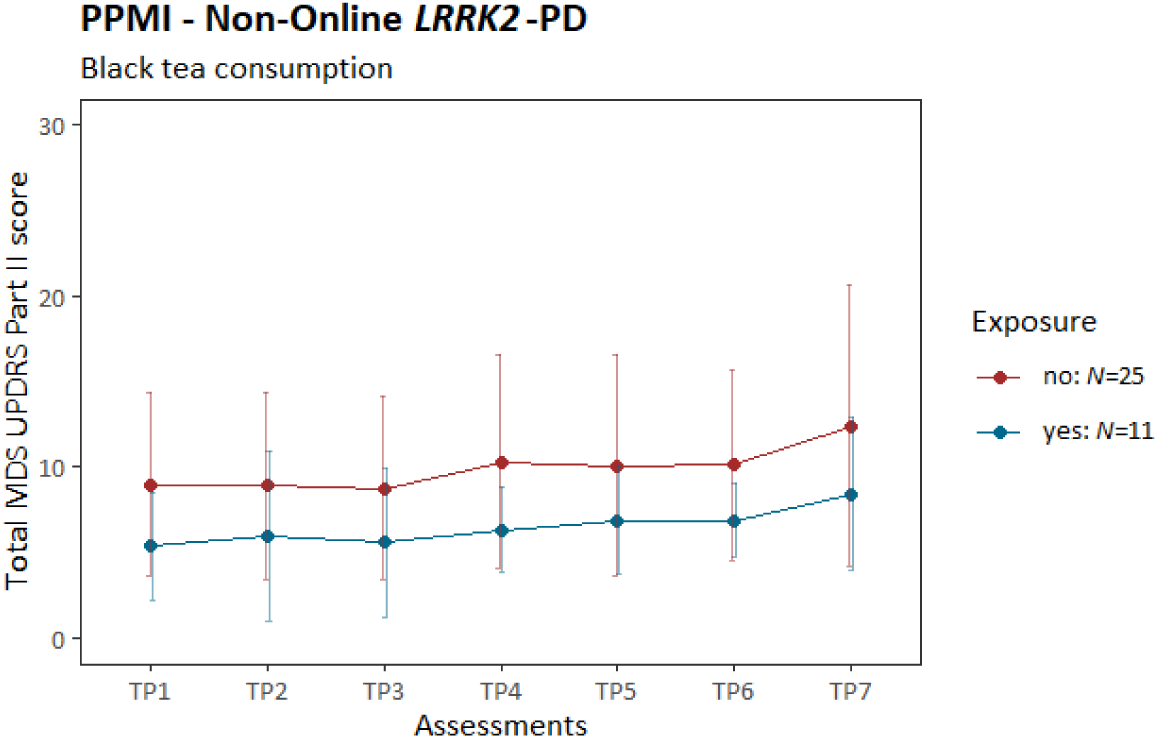
Motor signs severity over time stratified by black tea consumption. The plot shows the progression of PD motor features along the longitudinal assessments. The mean cumulative MDS-UPDRS Part II score is indicated at each time period, and the error bars show the corresponding standard deviation. Patients with *LRRK2*-PD (PPMI participants that are not enrolled in the PPMI-Online study) are shown. The patients are stratified by black tea consumption. *LRRK2*-PD=Patients with PD that carry the LRRK2 p.Gly2019Ser variant, TP=Time period, *N*=number of individuals.

**Supplementary Table 1.** The association between coffee consumption and motor sign severity over time. The motor sign severity was evaluated longitudinally over time and assessed with a mixed linear model in the PPMI and Fox Insight cohort.

|  | Estimate | SE | p-value |
| --- | --- | --- | --- |
| **PPMI – Online (iPD: yes=2059, no=548)** |  |  |  |
| Time period 2 | -0.002 | 0.01 | 0.844 |
| Time period 3 | 0.07 | 0.01 | $3.43 \times 10^{-12}$ |
| Time period 4 | 0.12 | 0.01 | $<2.00 \times 10^{-16}$ |
| Time period 5 | 0.20 | 0.01 | $<2.00 \times 10^{-16}$ |
| Time period 6 | 0.26 | 0.01 | $<2.00 \times 10^{-16}$ |
| Coffee | 0.05 | 0.03 | 0.157 |
| AAO | -0.06 | 0.01 | $8.97 \times 10^{-5}$ |
| Disease duration | 0.63 | 0.04 | $<2.00 \times 10^{-16}$ |
| **Fox Insight (iPD: yes=1054, no=312)** |  |  |  |
| Time period 2 | 0.02 | 0.01 | 0.218 |
| Time period 3 | 0.08 | 0.01 | $1.740 \times 10^{-8}$ |
| Time period 4 | 0.16 | 0.01 | $<2.00 \times 10^{-16}$ |
| Time period 5 | 0.23 | 0.02 | $<2.00 \times 10^{-16}$ |
| Time period 6 | 0.28 | 0.02 | $<2.00 \times 10^{-16}$ |
| Time period 7 | 0.32 | 0.02 | $<2.00 \times 10^{-16}$ |
| Coffee | 0.01 | 0.04 | 0.782 |
| AAO | -0.03 | 0.02 | 0.116 |
| Disease duration | 0.71 | 0.07 | $<2.00 \times 10^{-16}$ |
| OFF-episodes | 0.24 | 0.04 | $9.030 \times 10^{-11}$ |
| **Fox Insight (LRRK2-PD: yes=37, no=10)** |  |  |  |
| Time period 2 | 0.01 | 0.08 | 0.935 |
| Time period 3 | 0.06 | 0.08 | 0.404 |
| Time period 4 | 0.20 | 0.08 | 0.010 |
| Time period 5 | 0.18 | 0.08 | 0.020 |
| Time period 6 | 0.34 | 0.08 | $4.51 \times 10^{-5}$ |
| Time period 7 | 0.36 | 0.08 | $1.59 \times 10^{-5}$ |
| Coffee | -0.04 | 0.23 | 0.851 |
| AAO | -0.97 | 1.23 | 0.431 |
| Disease duration | 0.48 | 0.24 | 0.051 |
| OFF-episodes | 0.68 | 0.20 | 0.002 |

N=Number of individuals, iPD=idiopathic Parkinson's disease, LRRK2-PD= Patients with PD that carry the LRRK2 p.Gly2019Ser variant, Formula in R (package lme4): lmer(log(Cumulative MDS-UPDRS Part II Score) ~ Assessment time periods + mfp transformed AAO + mfp transformed disease duration + Environmental/Lifestyle factor (yes/no) + Experience of OFF episodes\* + (1|Patient ID))
\*If applicable
Baseline categories: Time period=Time period 1 (i.e., assessment at enrollment)

**Supplementary Table 2.**
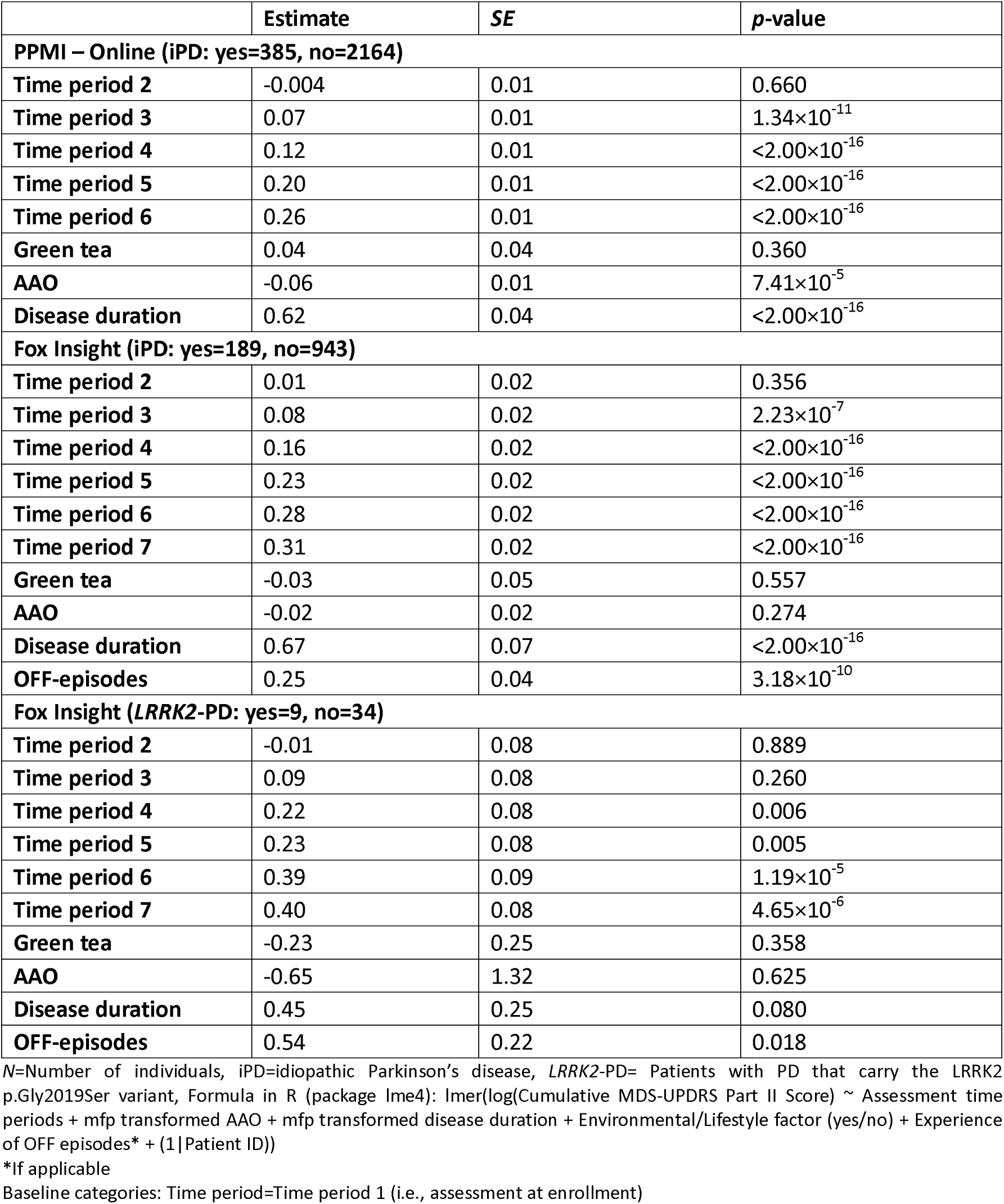
The association between green tea consumption and motor sign severity over time. The motor sign severity was evaluated longitudinally over time and assessed with a mixed linear model in the PPMI and Fox Insight cohort.

|  | Estimate | SE | p-value |
| --- | --- | --- | --- |
| **PPMI – Online (iPD: yes=385, no=2164)** |  |  |  |
| Time period 2 | -0.004 | 0.01 | 0.660 |
| Time period 3 | 0.07 | 0.01 | $1.34 \times 10^{-11}$ |
| Time period 4 | 0.12 | 0.01 | $<2.00 \times 10^{-16}$ |
| Time period 5 | 0.20 | 0.01 | $<2.00 \times 10^{-16}$ |
| Time period 6 | 0.26 | 0.01 | $<2.00 \times 10^{-16}$ |
| Green tea | 0.04 | 0.04 | 0.360 |
| AAO | -0.06 | 0.01 | $7.41 \times 10^{-5}$ |
| Disease duration | 0.62 | 0.04 | $<2.00 \times 10^{-16}$ |
| **Fox Insight (iPD: yes=189, no=943)** |  |  |  |
| Time period 2 | 0.01 | 0.02 | 0.356 |
| Time period 3 | 0.08 | 0.02 | $2.23 \times 10^{-7}$ |
| Time period 4 | 0.16 | 0.02 | $<2.00 \times 10^{-16}$ |
| Time period 5 | 0.23 | 0.02 | $<2.00 \times 10^{-16}$ |
| Time period 6 | 0.28 | 0.02 | $<2.00 \times 10^{-16}$ |
| Time period 7 | 0.31 | 0.02 | $<2.00 \times 10^{-16}$ |
| Green tea | -0.03 | 0.05 | 0.557 |
| AAO | -0.02 | 0.02 | 0.274 |
| Disease duration | 0.67 | 0.07 | $<2.00 \times 10^{-16}$ |
| OFF-episodes | 0.25 | 0.04 | $3.18 \times 10^{-10}$ |
| **Fox Insight (LRRK2-PD: yes=9, no=34)** |  |  |  |
| Time period 2 | -0.01 | 0.08 | 0.889 |
| Time period 3 | 0.09 | 0.08 | 0.260 |
| Time period 4 | 0.22 | 0.08 | 0.006 |
| Time period 5 | 0.23 | 0.08 | 0.005 |
| Time period 6 | 0.39 | 0.09 | $1.19 \times 10^{-5}$ |
| Time period 7 | 0.40 | 0.08 | $4.65 \times 10^{-6}$ |
| Green tea | -0.23 | 0.25 | 0.358 |
| AAO | -0.65 | 1.32 | 0.625 |
| Disease duration | 0.45 | 0.25 | 0.080 |
| OFF-episodes | 0.54 | 0.22 | 0.018 |

N=Number of individuals, iPD=idiopathic Parkinson's disease, LRRK2-PD= Patients with PD that carry the LRRK2 p.Gly2019Ser variant, Formula in R (package lme4): lmer(log(Cumulative MDS-UPDRS Part II Score) ~ Assessment time periods + mfp transformed AAO + mfp transformed disease duration + Environmental/Lifestyle factor (yes/no) + Experience of OFF episodes\* + (1 | Patient ID))
\*If applicable
Baseline categories: Time period=Time period 1 (i.e., assessment at enrollment)

**Supplementary Table 3.**
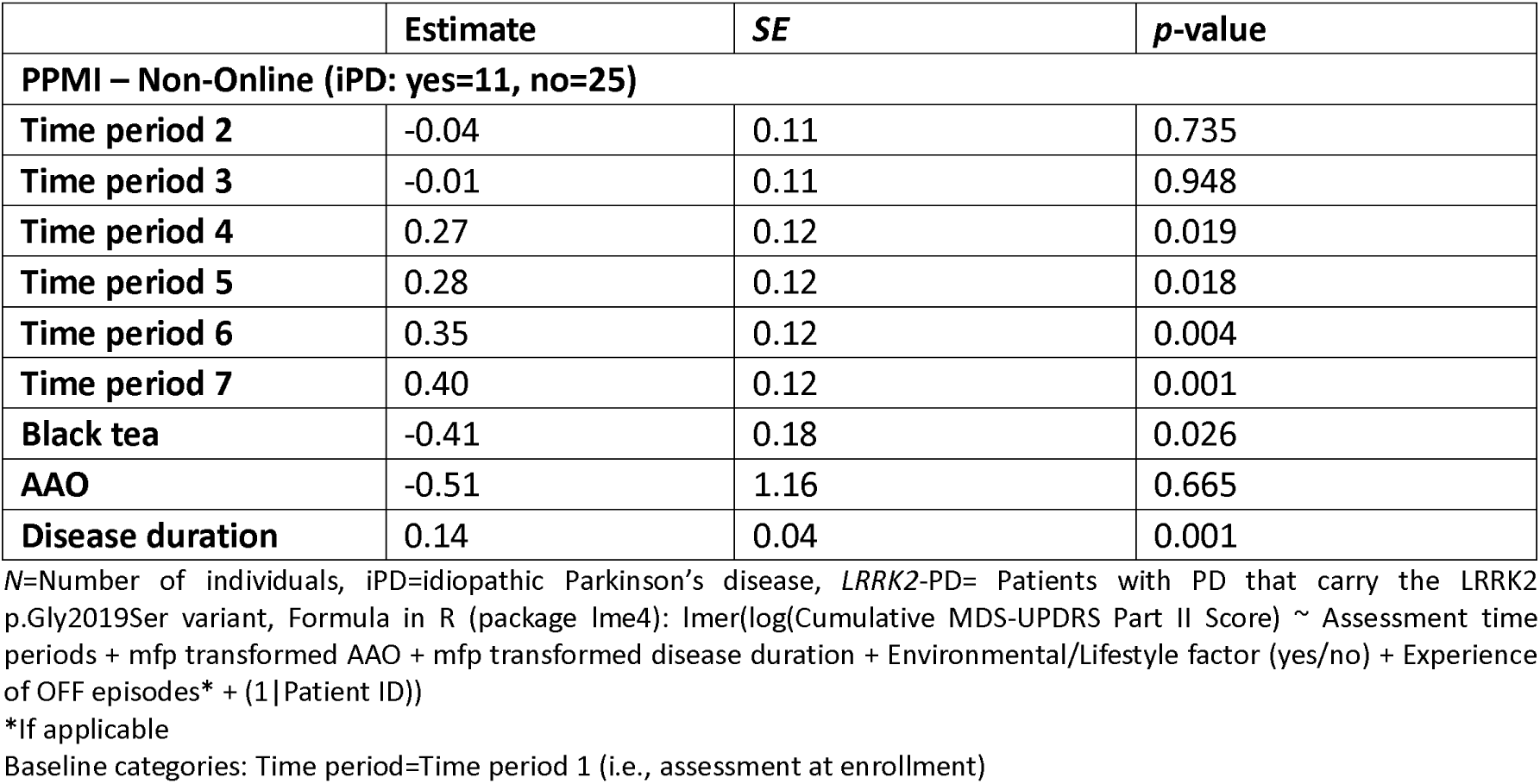
The association between black tea consumption and motor sign severity over time. The motor sign severity was evaluated longitudinally over time and assessed with a mixed linear model in patients with *LRRK2*-PD PPMI participants who are not enrolled in the PPMI-Online study.

