## Supplementary for "Pesticides and lifestyle factors are associated with disease severity of Parkinson’s disease: a longitudinal study"

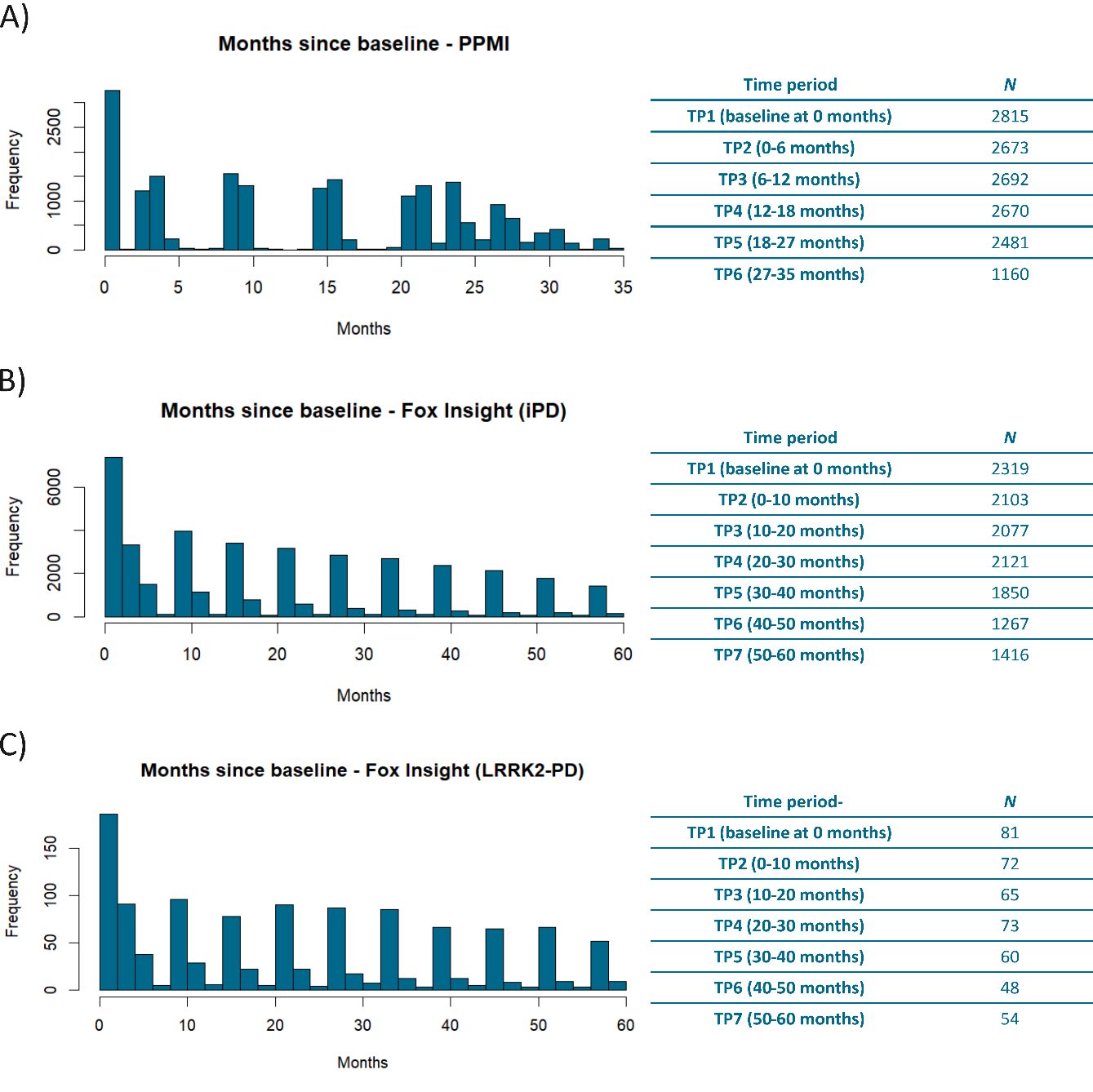
**Supplementary Material:**

**Supplementary Figure 1. Enrollment of Patients in the PPMI-Online and Fox Insight longitudinal cohort.**

The bar charts show the number of patients with PD that longitudinally responded to the motor signs assessment in the months since baseline. The assessments were stratified into six time periods (PPMI, **A**) or seven (Fox Insight iPD and LRRK2-PD, **B-C**). The number of patients per assessment is also presented in the corresponding tables. *N*=Number of individuals, iPD=idiopathic Parkinson’s disease, *LRRK2*-PD= Patients with PD that carry the LRRK2 p.Gly2019Ser variant.

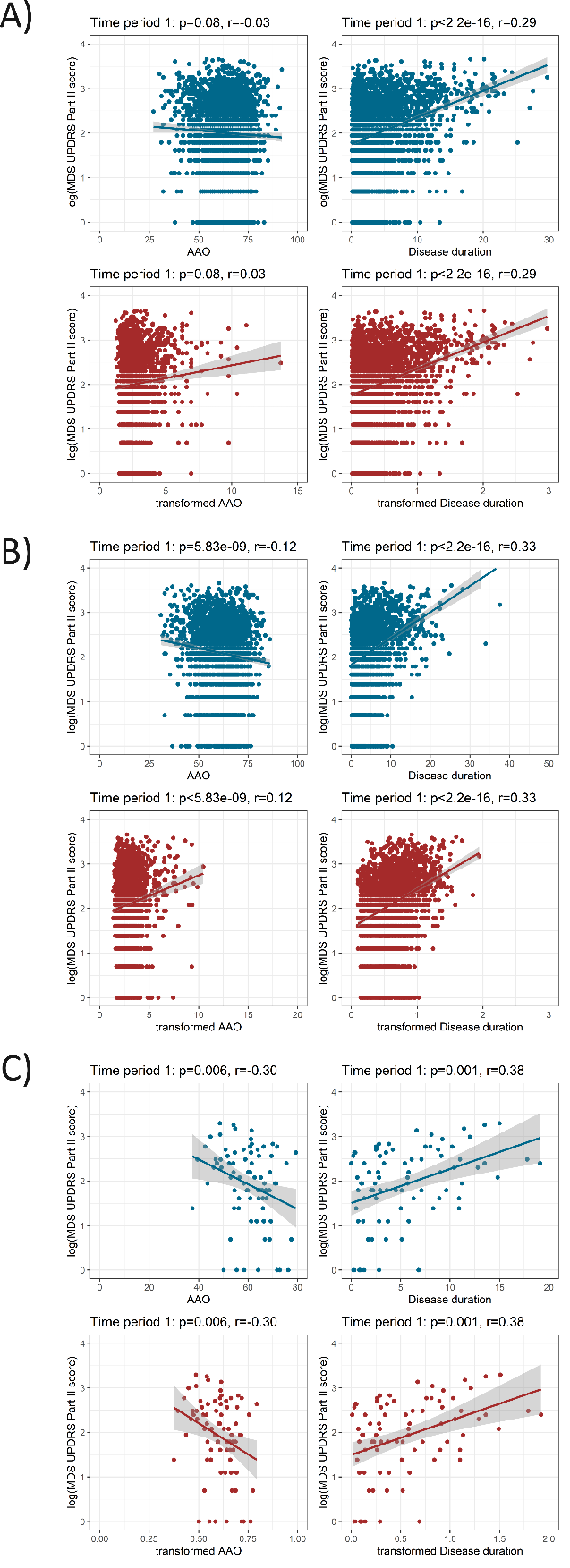

**Supplementary Figure 2. Association between age at onset or disease duration and motor signs score.**

The scatter plots show the relationship between the age of disease onset (AAO) and disease duration with the logarithmized MDS-UPDRS part II score. The untransformed data are shown in blue, and the transformed data in red (by using fractional polynomials). The plots show the data of patients with iPD (PPMI-Online **A** and Fox Insight in **B**) or with LRRK2-PD (Fox Insight in **C**). iPD=idiopathic Parkinson’s disease, *LRRK2*-PD=Patients with PD that carry the LRRK2 p.Gly2019Ser variant, *r*=Spearman's rank correlation rho, *p*=Spearman's rank correlation *p*-value.

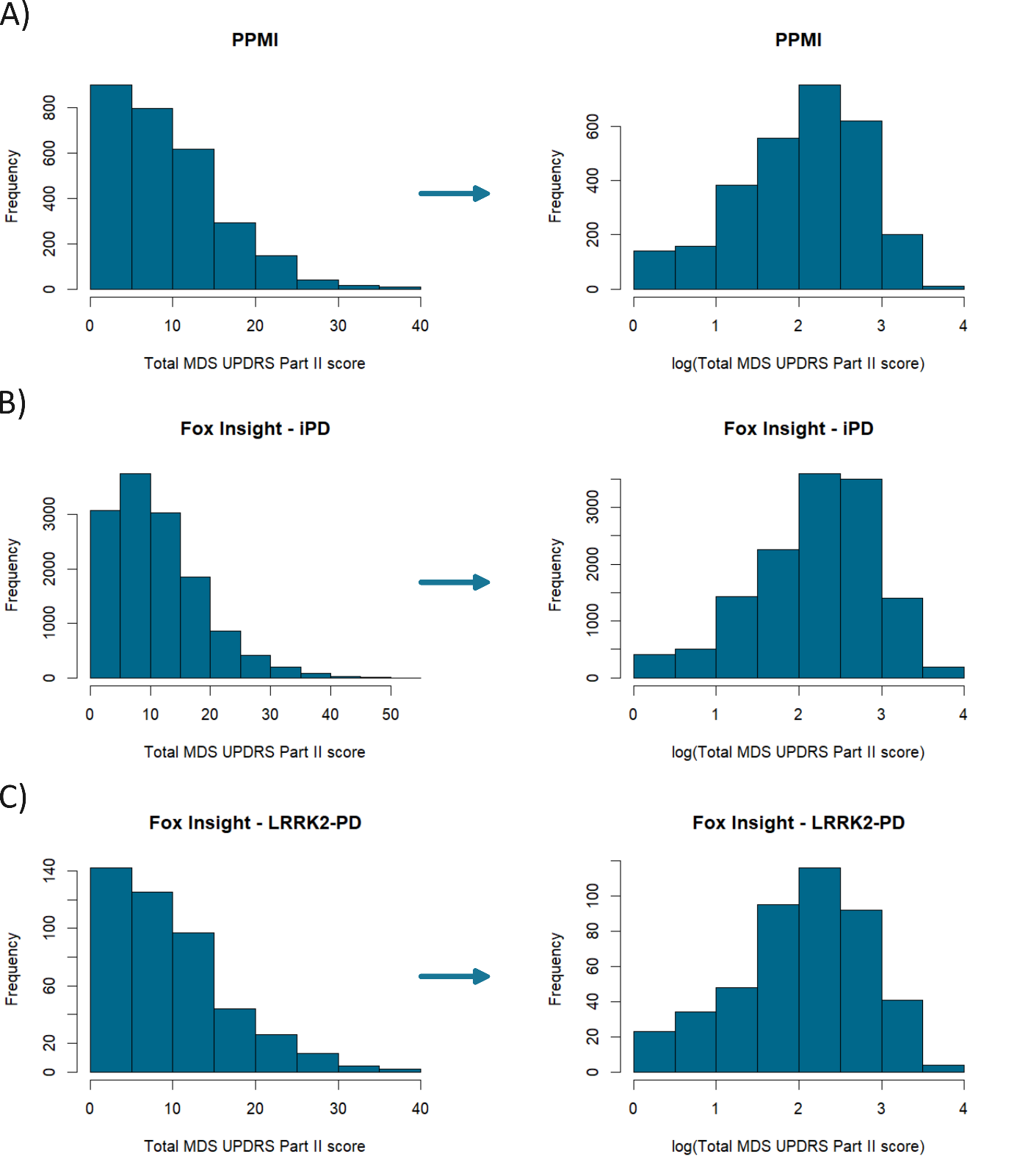

**Supplementary Figure 3. Distribution of the motor signs severity score.**

The histograms show the frequency of the cumulative MDS-UPDRS Part II score. As there is no normal distribution the score has been logarithmized. The plots show the data of patients with iPD (PPMI-Online **A** and Fox Insight in **B**) or with LRRK2-PD (Fox Insight in **C**). iPD=idiopathic Parkinson’s disease, *LRRK2*-PD=Patients with PD that carry the LRRK2 p.Gly2019Ser variant.

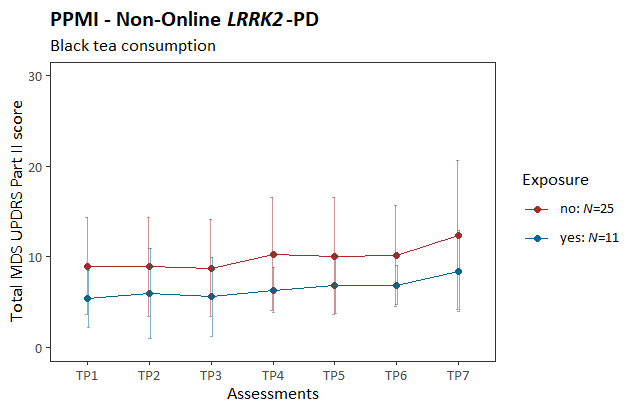

**Supplementary Figure 4. Motor signs severity over time stratified by black tea consumption.**

The plot shows the progression of PD motor features along the longitudinal assessments. The mean cumulative MDS-UPDRS Part II score is indicated at each time period, and the error bars show the corresponding standard deviation. Patients with *LRRK2*-PD (PPMI participants that are not enrolled in the PPMI-Online study) are shown. The patients are stratified by black tea consumption. *LRRK2*-PD=Patients with PD that carry the LRRK2 p.Gly2019Ser variant, TP=Time period, *N*=number of individuals.

**Supplementary Table 1.** The association between coffee consumption and motor sign severity over time. The motor sign severity was evaluated longitudinally over time and assessed with a mixed linear model in the PPMI and Fox Insight cohort.

|  | **Estimate** | ***SE*** | ***p*-value** |
| --- | --- | --- | --- |
| **PPMI – Online (iPD: yes=2059, no=548)** | | | |
| **Time period 2** | -0.002 | 0.01 | 0.844 |
| **Time period 3** | 0.07 | 0.01 | 3.43×10^-12^ |
| **Time period 4** | 0.12 | 0.01 | <2.00×10^-16^ |
| **Time period 5** | 0.20 | 0.01 | <2.00×10^-16^ |
| **Time period 6** | 0.26 | 0.01 | <2.00×10^-16^ |
| **Coffee** | 0.05 | 0.03 | 0.157 |
| **AAO** | -0.06 | 0.01 | 8.97×10^-5^ |
| **Disease duration** | 0.63 | 0.04 | <2.00×10^-16^ |
| **Fox Insight (iPD: yes=1054, no=312)** | | | |
| **Time period 2** | 0.02 | 0.01 | 0.218 |
| **Time period 3** | 0.08 | 0.01 | 1.740×10^-8^ |
| **Time period 4** | 0.16 | 0.01 | <2.00×10^-16^ |
| **Time period 5** | 0.23 | 0.02 | <2.00×10^-16^ |
| **Time period 6** | 0.28 | 0.02 | <2.00×10^-16^ |
| **Time period 7** | 0.32 | 0.02 | <2.00×10^-16^ |
| **Coffee** | 0.01 | 0.04 | 0.782 |
| **AAO** | -0.03 | 0.02 | 0.116 |
| **Disease duration** | 0.71 | 0.07 | <2.00×10^-16^ |
| **OFF-episodes** | 0.24 | 0.04 | 9.030×10^-11^ |
| **Fox Insight (*LRRK2*-PD: yes=37, no=10)** | | | |
| **Time period 2** | 0.01 | 0.08 | 0.935 |
| **Time period 3** | 0.06 | 0.08 | 0.404 |
| **Time period 4** | 0.20 | 0.08 | 0.010 |
| **Time period 5** | 0.18 | 0.08 | 0.020 |
| **Time period 6** | 0.34 | 0.08 | 4.51×10^-5^ |
| **Time period 7** | 0.36 | 0.08 | 1.59×10^-5^ |
| **Coffee** | -0.04 | 0.23 | 0.851 |
| **AAO** | -0.97 | 1.23 | 0.431 |
| **Disease duration** | 0.48 | 0.24 | 0.051 |
| **OFF-episodes** | 0.68 | 0.20 | 0.002 |

*N*=Number of individuals, iPD=idiopathic Parkinson’s disease, *LRRK2*-PD= Patients with PD that carry the LRRK2 p.Gly2019Ser variant, Formula in R (package lme4): lmer(log(Cumulative MDS-UPDRS Part II Score) ~ Assessment time periods + mfp transformed AAO + mfp transformed disease duration + Environmental/Lifestyle factor (yes/no) + Experience of OFF episodes* + (1|Patient ID))

*If applicable

Baseline categories: Time period=Time period 1 (i.e., assessment at enrollment)

**Supplementary Table 2.** The association between green tea consumption and motor sign severity over time. The motor sign severity was evaluated longitudinally over time and assessed with a mixed linear model in the PPMI and Fox Insight cohort.

|  | **Estimate** | ***SE*** | ***p*-value** |
| --- | --- | --- | --- |
| **PPMI – Online (iPD: yes=385, no=2164)** | | | |
| **Time period 2** | -0.004 | 0.01 | 0.660 |
| **Time period 3** | 0.07 | 0.01 | 1.34×10^-11^ |
| **Time period 4** | 0.12 | 0.01 | <2.00×10^-16^ |
| **Time period 5** | 0.20 | 0.01 | <2.00×10^-16^ |
| **Time period 6** | 0.26 | 0.01 | <2.00×10^-16^ |
| **Green tea** | 0.04 | 0.04 | 0.360 |
| **AAO** | -0.06 | 0.01 | 7.41×10^-5^ |
| **Disease duration** | 0.62 | 0.04 | <2.00×10^-16^ |
| **Fox Insight (iPD: yes=189, no=943)** | | | |
| **Time period 2** | 0.01 | 0.02 | 0.356 |
| **Time period 3** | 0.08 | 0.02 | 2.23×10^-7^ |
| **Time period 4** | 0.16 | 0.02 | <2.00×10^-16^ |
| **Time period 5** | 0.23 | 0.02 | <2.00×10^-16^ |
| **Time period 6** | 0.28 | 0.02 | <2.00×10^-16^ |
| **Time period 7** | 0.31 | 0.02 | <2.00×10^-16^ |
| **Green tea** | -0.03 | 0.05 | 0.557 |
| **AAO** | -0.02 | 0.02 | 0.274 |
| **Disease duration** | 0.67 | 0.07 | <2.00×10^-16^ |
| **OFF-episodes** | 0.25 | 0.04 | 3.18×10^-10^ |
| **Fox Insight (*LRRK2*-PD: yes=9, no=34)** | | | |
| **Time period 2** | -0.01 | 0.08 | 0.889 |
| **Time period 3** | 0.09 | 0.08 | 0.260 |
| **Time period 4** | 0.22 | 0.08 | 0.006 |
| **Time period 5** | 0.23 | 0.08 | 0.005 |
| **Time period 6** | 0.39 | 0.09 | 1.19×10^-5^ |
| **Time period 7** | 0.40 | 0.08 | 4.65×10^-6^ |
| **Green tea** | -0.23 | 0.25 | 0.358 |
| **AAO** | -0.65 | 1.32 | 0.625 |
| **Disease duration** | 0.45 | 0.25 | 0.080 |
| **OFF-episodes** | 0.54 | 0.22 | 0.018 |

*N*=Number of individuals, iPD=idiopathic Parkinson’s disease, *LRRK2*-PD= Patients with PD that carry the LRRK2 p.Gly2019Ser variant, Formula in R (package lme4): lmer(log(Cumulative MDS-UPDRS Part II Score) ~ Assessment time periods + mfp transformed AAO + mfp transformed disease duration + Environmental/Lifestyle factor (yes/no) + Experience of OFF episodes* + (1|Patient ID))

*If applicable

Baseline categories: Time period=Time period 1 (i.e., assessment at enrollment)

**Supplementary Table 3.** The association between black tea consumption and motor sign severity over time. The motor sign severity was evaluated longitudinally over time and assessed with a mixed linear model in patients with *LRRK2*-PD PPMI participants who are not enrolled in the PPMI-Online study.

|  | **Estimate** | ***SE*** | ***p*-value** |
| --- | --- | --- | --- |
| **PPMI – Non-Online (iPD: yes=11, no=25)** | | | |
| **Time period 2** | -0.04 | 0.11 | 0.735 |
| **Time period 3** | -0.01 | 0.11 | 0.948 |
| **Time period 4** | 0.27 | 0.12 | 0.019 |
| **Time period 5** | 0.28 | 0.12 | 0.018 |
| **Time period 6** | 0.35 | 0.12 | 0.004 |
| **Time period 7** | 0.40 | 0.12 | 0.001 |
| **Black tea** | -0.41 | 0.18 | 0.026 |
| **AAO** | -0.51 | 1.16 | 0.665 |
| **Disease duration** | 0.14 | 0.04 | 0.001 |

*N*=Number of individuals, iPD=idiopathic Parkinson’s disease, *LRRK2*-PD= Patients with PD that carry the LRRK2 p.Gly2019Ser variant, Formula in R (package lme4): lmer(log(Cumulative MDS-UPDRS Part II Score) ~ Assessment time periods + mfp transformed AAO + mfp transformed disease duration + Environmental/Lifestyle factor (yes/no) + Experience of OFF episodes* + (1|Patient ID))

*If applicable

Baseline categories: Time period=Time period 1 (i.e., assessment at enrollment)
